# Transmission heterogeneities, kinetics, and controllability of SARS-CoV-2

**DOI:** 10.1101/2020.08.09.20171132

**Authors:** Kaiyuan Sun, Wei Wang, Lidong Gao, Yan Wang, Kaiwei Luo, Lingshuang Ren, Zhifei Zhan, Xinghui Chen, Shanlu Zhao, Yiwei Huang, Qianlai Sun, Ziyan Liu, Maria Litvinova, Alessandro Vespignani, Marco Ajelli, Cécile Viboud, Hongjie Yu

## Abstract

A long-standing question in infectious disease dynamics concerns the role of transmission heterogeneities, driven by demography, behavior and interventions. Based on detailed patient and contact tracing data in Hunan, China we find 80% of secondary infections traced back to 15% of SARS-CoV-2 primary infections, indicating substantial transmission heterogeneities. Transmission risk scales positively with the duration of exposure and the closeness of social interactions and is modulated by demographic and clinical factors. The lockdown period increases transmission risk in the family and households, while isolation and quarantine reduce risks across all types of contacts. The reconstructed infectiousness profile of a typical SARS-CoV-2 patient peaks just before symptom presentation. Modeling indicates SARS-CoV-2 control requires the synergistic efforts of case isolation, contact quarantine, and population-level interventions, owing to the specific transmission kinetics of this virus.

**One Sentence Summary:** Public health measures to control SARS-CoV-2 could be designed to block the specific transmission characteristics of the virus.

## Main Text

While it has been well documented that the clinical severity of COVID-19 increases with age (*1*–*5*), information is limited on how transmission risk varies with demographic factors, clinical presentation, and contact type (*6*–*12*). Individual-based interventions such as case isolation, contact tracing and quarantine have been shown to accelerate case detection and interrupt transmission chains (*13*). However, these interventions are typically implemented in conjunction with population-level physical distancing measures, and their effect on contact patterns and transmission risk remains difficult to separate (*14*–*24*). A better understanding of the factors driving SARS-CoV-2 transmission is key to achieve epidemic control while minimizing societal cost, particularly as countries relax physical distancing measures.

Hunan, a province in China adjacent to Hubei where the COVID-19 pandemic began, experienced sustained SARS-CoV-2 transmission in late January and early February 2020, followed by a quick suppression of the outbreak by March 2020. As in many other provinces in China, epidemic control was achieved by layering interventions targeting SARS-CoV-2 cases and their contacts with population-level physical distancing measures. In this study, we reconstruct transmission chains among all identified SARS-CoV-2 infections in Hunan, as of April 3, 2020, based on granular epidemiological information collected through extensive surveillance and contact tracing efforts. We identify the demographic, clinical and behavioral factors that drive transmission heterogeneities and evaluate how interventions modulate the topology of the transmission network. Further, we reconstruct the infectiousness profile of SARS-CoV-2 over the course of a typical infection and estimate the feasibility of epidemic control by individual and population-based interventions.

We analyze detailed epidemiological records for 1,178 SARS-CoV-2 infected individuals and their 15,648 close contacts, representing 19,227 separate exposure events, compiled by the Hunan Provincial Center for Disease Control and Prevention. Cases were identified between January 16 and April 3, 2020; primary cases were captured by passive surveillance, contact tracing or travel screening and were laboratory confirmed by RT-PCR. Individuals who were close contacts of the primary cases were followed for at least 2 weeks after the last exposure to the infected individual. Prior to February 7, 2020, contacts were tested if they developed symptoms during the quarantine period. After February 7, 2020, RT-PCR testing was required for all contacts, and specimens were collected at least once from each contact during quarantine, regardless of symptoms. Upon positive RT-PCR test results, infected individuals were isolated in dedicated hospitals, regardless of their clinical severity, while their contacts were quarantined in medical observation facilities. The case ascertainment process is visualized in Fig S1.

The dataset includes 210 epidemiological clusters representing 831 cases, with additional 347 sporadic cases (29%) unlinked to any cluster (see Supplementary Materials & Methods for more details). For each cluster, we stochastically reconstruct transmission chains and estimate the timing of infection most compatible with each patient’s exposure history. We analyze an ensemble of 100 reconstructed transmission chains to account for uncertainties in exposure histories (Fig. 1 visualizes one realization of the transmission chains, while Fig S2A illustrates variability in the topology of the aggregation of 100 realizations transmission chains).

**Fig. 1.**
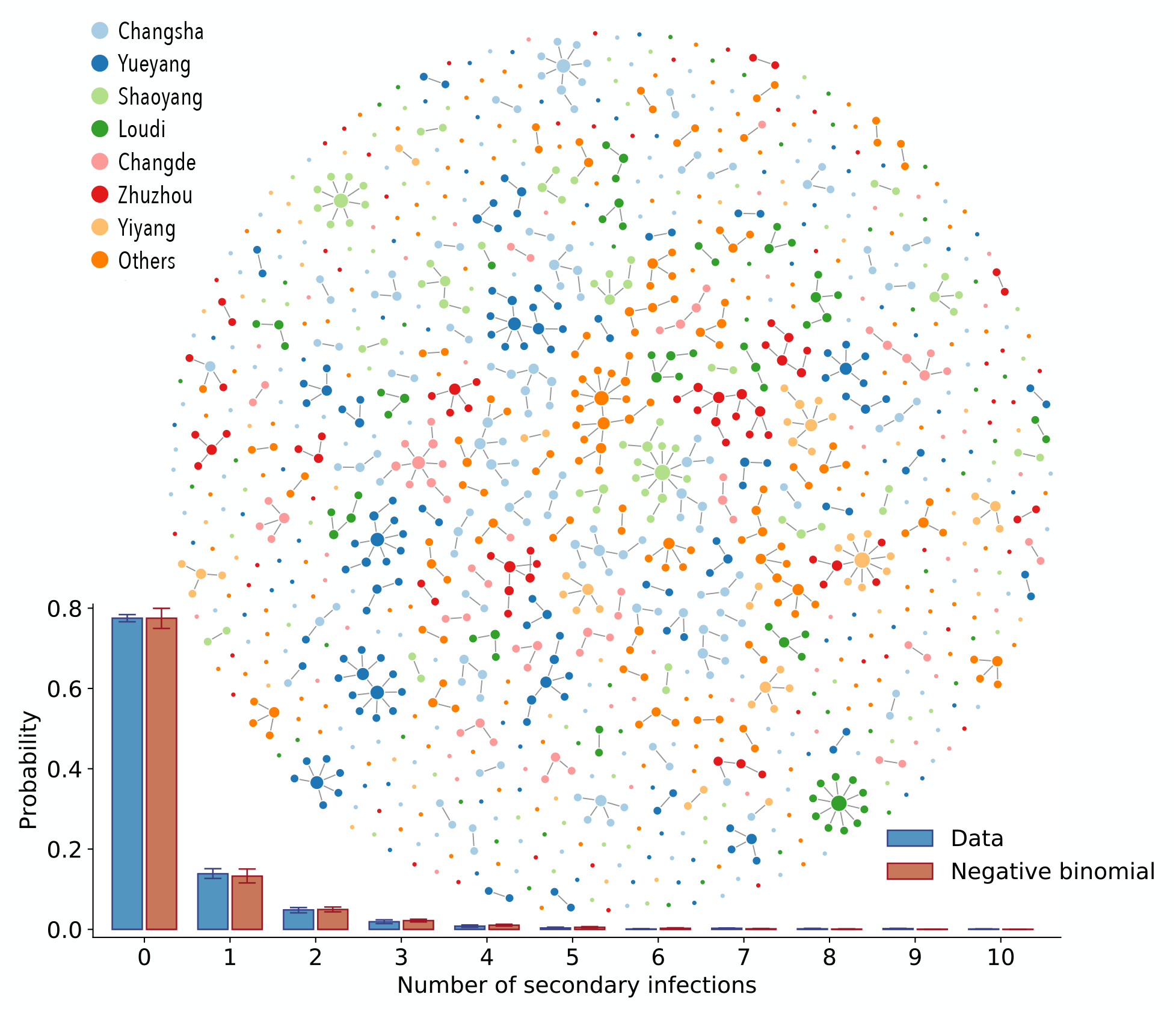
SARS-CoV-2 transmission chains. Top: One realization of the reconstructed transmission chains among 1,178 SARS-CoV-2 infected individuals in Hunan province. Each node in the network represents a patient infected with SARS-CoV-2 and each link represents an infector-infectee relationship. The color of the node denotes the reporting prefecture of infected individuals. Bottom: Distribution of the number of secondary infections; blue bars represent the ensemble averaged across 100 stochastic samples of the reconstructed transmission chains. Orange bars represent the best fit of a negative binomial distribution to the ensemble average. Vertical lines indicate 95% confidence intervals across 100 samples (of both data and the models’ fitting results). Some confidence intervals are narrow and not visible on the plot. For sensitivity analysis, we also fit the distribution with geometric and Poisson distribution. Based on the Akaike information criterion, the negative binomial distribution fit the data the best (average AIC score for negative binomial distribution: 1902; for geometric distribution: 1981, for Poisson distribution: 2259).

We observe between 0 and 4 generations of transmission, with the largest cluster involving 20 SARS-CoV-2-infected individuals. The number of secondary infections ranges from 0 to 10, with a distribution of secondary infections best characterized by a negative binomial distribution with mean µ = 0.40 (95% CI, 0.35 to 0.47) and variance µ(1 + µ/k) = 0.96 (95% CI, 0.74 to 1.26), where *k* = 0.30 (95% CI, 0.23 to 0.39) is the dispersion parameter (Fig. 1). We find that 80% of secondary infections can be traced back to 15% of SARS-CoV-2 infected individuals, indicating substantial transmission heterogeneities at the individual-level. We can also assess geographic diffusion within Hunan province and find that the great majority of transmission events occur within the same prefecture (94.3%, 95%CI, 93.7% to 95.0%), with occasional spread between prefectures (5.7%, 95%CI, 5.0% to 6.3%).

### Characterizing SARS-CoV-2 transmission heterogeneities at the individual level

To dissect the individual transmission heterogeneities and identify predictors of transmission, we analyze the infection risk among a subset of 14,622 individuals who were close contacts of 870 SARS-CoV-2 patients. This dataset excludes primary cases whose infected contacts report a travel history to Wuhan. The dataset represents 74% of all SARS-CoV-2 cases recorded in the Hunan patient database. Contacts of these 870 patients have been carefully monitored, so that 17,750 independent exposure events have been captured.

We start by characterizing variation in transmission risk across the diverse set of 17,750 exposures. We study how the per-contact transmission risk varies with the type of exposures, exposure duration, exposure timing, and physical distancing intervention, after adjusting for demographic, clinical, and travel-related factors. Exposures are grouped into 5 categories based on contact type, namely: household, extended family, social, community, and healthcare (Table S2), with the duration of exposure approximated by the time interval between the initial and final dates of exposure. To gauge the impact of physical distancing on transmission risks, we further stratify exposures by the date of occurrence, with January 25, 2020 marking the beginning of lockdown in Hunan (based on Baidu Qianxi mobility index (*25*), Fig. S3A insert). To address putative variation in infectiousness over the course of infection, we distinguish whether exposures overlap with the date of symptom onset of a primary case, a period associated with high viral shedding. We use a mixed effects multiple logistic regression model (GLMM-logit) to quantify the effects of these factors on the per-contact risk of transmission (see Table S3 for a detailed definition of all risk factors and summary statistics).

Based on the point estimates of the regression (see Fig. S3A for regression results), we find that household contacts pose the highest risk of transmission, followed by extended family, social and community contacts, in agreement with a prior study (*12*). Healthcare contacts have the lowest risk, suggesting that adequate protective measures were adopted by patients and healthcare staff in Hunan. Interestingly, the impact of physical distancing differs by contact type (Table 1): the risk of transmission in the household increases during the lockdown period, likely due to increased contact frequency at home as a result of physical confinement. In contrast, the transmission risk decreases for community and social contacts during lockdown, possibly due to adoption of prudent behaviors such as mask wearing, hand washing and coughing/sneezing etiquette. We find that longer exposures are riskier, with one additional day of exposure increasing the transmission risk by 10% (95% CI, 5% to 15%). Further, transmission risk is higher around the time of symptom presentation of the primary case (Table 1). In addition, susceptibility to infection (defined as the risk of infection given a contact with primary case) by age: children aged 0-12 years are significantly less susceptible than individuals 26-64 years (odds ratio 0.41, 95% CI 0.26 to 0.63); while patients older than 65 years are significantly more susceptible (odds ratio 1.39, 95% CI 1.02 to 1.91). In contrast, we find no statistical support for age difference in infectivity (Fig. S3A). These results are in agreement with previous findings (*12, 26, 27*).

**Table 1:** SARS-CoV-2 transmission risk in Hunan by contact type, duration of exposure, and whether the exposure window contains the date of symptom onset of the primary case - a period of intense viral shedding. Risk is further stratified by the date of implementation of social distancing interventions in Hunan, which is 01/25/2020. The regression model is adjusted for demographic characteristics of the cases and their contacts, clinical symptoms, and travel history. Details are provided in the Material and Methods, while the full results of the regression including additional risk factors are shown in Fig. S3. * indicates p-value<0.05, ** indicates p-value<0.01, *** indicates p-value<0.001.

| Risk factors |  | Odds ratio | 95% CI |
| --- | --- | --- | --- |
| Household contacts | Before 01/25/2020 | 2.20*** | (1.39, 3.49) |
|  | After 01/25/2020 | 3.79*** | (2.47, 5.79) |
| Extended Family contacts | Before 01/25/2020 | 1.00 | Reference |
|  | After 01/25/2020 | 0.94 | (0.60, 1.46) |
| Social contacts | Before 01/25/2020 | 0.63 | (0.37, 1.06) |
|  | After 01/25/2020 | 0.41** | (0.21, 0.78) |
| Community contacts | Before 01/25/2020 | 0.37** | (0.19, 0.74) |
|  | After 01/25/2020 | 0.2* | (0.05, 0.71) |
| Healthcare contacts | Before 01/25/2020 | 0.15* | (0.03, 0.68) |
|  | After 01/25/2020 | 0.10* | (0.01, 0.90) |
| Duration of exposure (days) |  | 1.10*** | (1.05, 1.15) |
| Symptom onset within exposure window (Yes) |  | 1.49* | (1.09, 2.04) |

For each of the 17,750 contact exposure events, we estimate the probability of transmission using the point estimate of the baseline odds and the odds ratios from the GLMM-logit regression (Fig. S3A). In Fig. 2A, we plot the distribution of transmission probabilities for household, extended family, social, and community contacts separately. The average per-contact transmission probability is highest for household contacts (7.2%, 95% CI, 1.2% to 19.6%), followed by family (1.7%, 95% CI, 0.4% to 5.6%) and social contacts (0.9%, 95% CI, 0.2% to 2.7%); while the risk is lowest for community contacts (0.4%, 95% CI, 0.1% to 1.1%). These transmission probabilities reflect the joint effect of duration of exposure (Fig. 2B), superimposed on differences in transmission risk by type of contact (Fig. S3A). While confidence intervals on risk estimates are broad, there is statistical support for separating out contacts in 5 categories and including a time covariate to capture the effect of the lockdown, rather than collapsing the contact data into fewer categories (Table S4). In contrast, there is no statistical support for a more complex model that considers a different effect of contact duration by type of contact (Table S4). It is worth noting that the per-contact transmission probabilities were estimated in a situation of intense interventions and high population awareness of the disease, and thus, they may be not generalizable elsewhere.

**Fig. 2.**
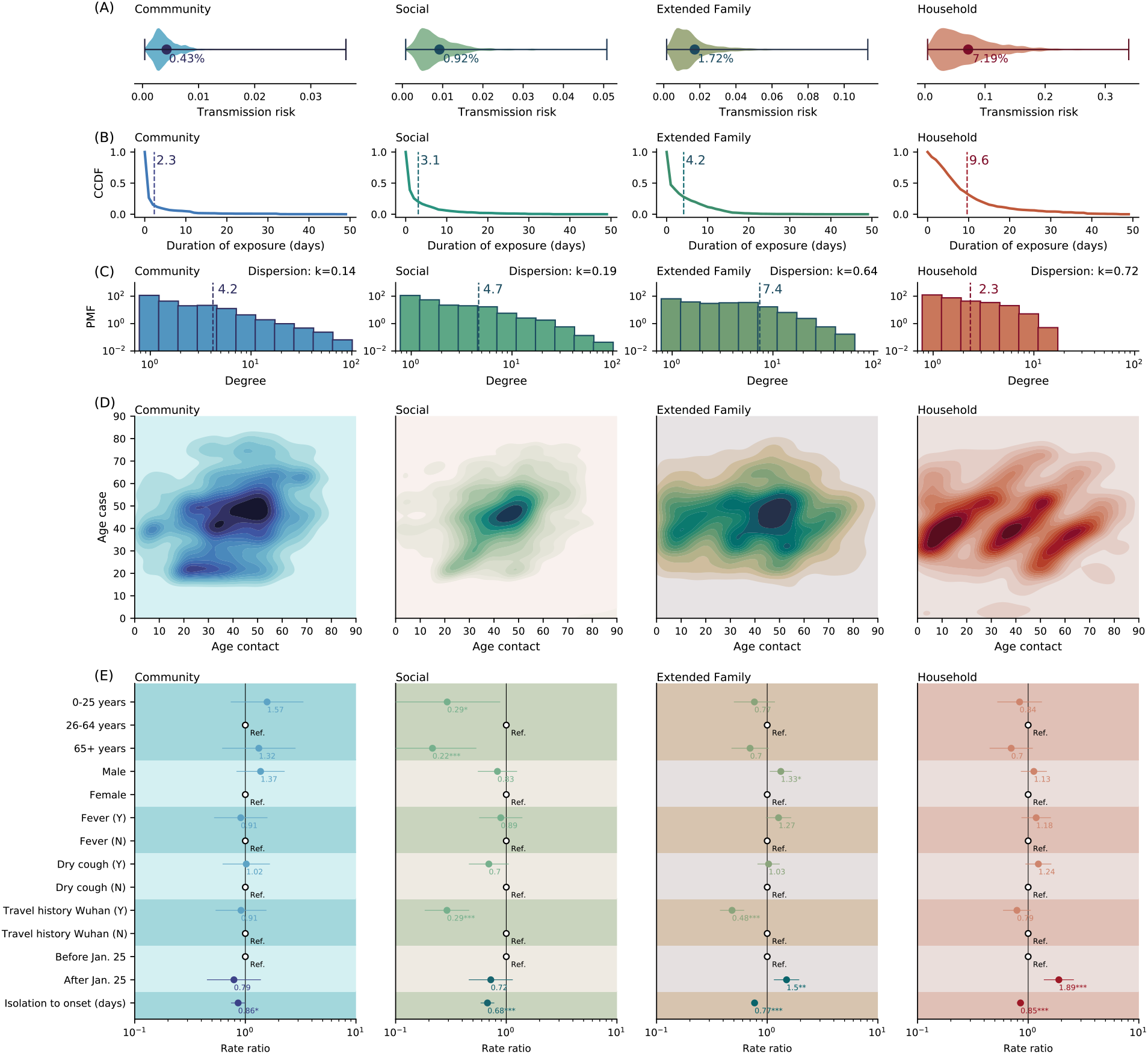
Heterogeneity in contact rates of SARS-CoV-2 cases and impact of interventions, by contact type. Columns from left to right represent community contacts (public transportation, food & entertainment), social contacts, extended family contacts, and household contacts. (A) Violin plots representing the distribution of per-contact transmission probability by contact type, adjusted for all other covariates in Fig. S3 (probability expressed in percentage, x-axis). (B) Complementary cumulative distribution function (CCDF, y axis) for duration of exposure (i.e. the probability that exposure is longer or equal to a certain value). Dashed vertical lines indicate average values. Household contacts last the longest, and as expected contact duration decreases as social ties loosen. (C) The distribution of the number of unique contacts (degree distribution) of the primary cases for each contact types. PMF on the y axis is acronym for probability mass function. The dashed vertical lines indicate average values. The dispersion parameter *k* is calculated based on the relationship 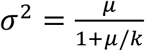, where *μ* and *σ*^2^ are mean and variance of the number of unique contacts. *k* < 1 indicates over-dispersion. (D) Age distribution of SARS-CoV-2 case-contact pairs (contact matrices). (E) Rate ratios of negative binomial regression of the cumulative contact rates (CCRs) against predictors including the infector’s age, sex, presence of fever/cough, Wuhan travel history, whether symptom onset occurred before social distancing was in place (before or after Jan. 25, 2020), and time from isolation to symptom onset. CCRs represent the sum of relevant contacts over a one-week window centered at the date of the primary case’s symptom onset. Dots and lines indicate point estimates and 95% confidence interval of the rate ratios, numbers below the dots indicate the numerical value of the point estimates; Ref. stands for reference category; * indicates p-value<0.05, ** indicates p-value<0.01, *** indicates p-value<0.001.

The number of contacts is also a key driver of individual transmission potential and varies by contact type. Fig. 2C presents the contact degree distribution, defined as the number of unique contacts per individual. We find that the distributions of individual contact degree are over-dispersed with dispersion parameter 0 < *k* < 1 across all contact types. Furthermore, household (*k* = 0.72) and extended family (*k* = 0.64) contacts are less dispersed than social (*k* = 0.19) and community (*k*= 0.14) contacts, suggesting that contact heterogeneities are inversely correlated with the closeness of social interactions. Fig. 2D visualizes the age-specific contact patterns between the primary cases and their contacts, demonstrating diverse mixing patterns across different types of contact. Specifically, household contacts present the canonical “three-bands” pattern where the diagonal illustrates age-assortative interactions and the two off-diagonals represent inter-generational mixing (*28, 29*). Other contact types display more diffusive mixing patterns by age. We also see that among all primary cases, young and middle-aged adults have the most social contacts (Fig 2E).

Next, we summarize the overall transmission potential of an individual by calculating the cumulative contact rate (CCR) of all primary cases. The CCR captures how contact opportunities vary with demography, temporal variation in the infectiousness profile, an individual’s contact degree, and interventions. (See Section 4.3 in Materials and Methods for detailed definition). After adjusting for age, sex, clinical presentation, and travel history to Wuhan, we find that physical distancing measures increase CCRs for household and extended family contacts and decrease (although not statistically significant) CCRs for social and community contacts (Fig. 2E). In contrast, faster case isolation universally reduces CCRs, decreasing transmission opportunities across all contact types (Fig. 2E).

### Characterizing the natural history of SARS-CoV-2 infection by strength of interventions

We have characterized SARS-CoV-2 transmission risk factors and have shown that individual and population-based interventions have a differential impact on contact patterns and transmission potential. Next, we use our probabilistic reconstruction of infector-infectee pairs to further dissect transmission kinetics and project the impact of interventions on SARS-CoV-2 dynamics. Based on the reconstructed transmission chains, we estimate a median serial interval of 5.3 days, with an inter-quartile range (IQR) of 2.7 to 8.3 days, which represents the time interval between symptom onset of an infector and his/her infectee (Fig. S7B, D). The median generation interval, defined as the interval between the infection times of an infector and his/her infectee, is 5.3 days, with an IQR of 3.1 to 8.7 days (Fig. S7A, C). We estimate that 63.4% (95% CI, 60.2% to 67.2%) of all transmission events occur before symptom onset, which is comparable with findings from other studies (*6–8, 10–13, 19, 30, 31*). However, these estimates are impacted by the intensity of interventions; in Hunan, isolation and quarantine were in place throughout the epidemic.

Case isolation and contact quarantine are meant to prevent potentially infectious individuals from contacting susceptible individuals, effectively shortening the infectious period. As a result, we would expect right censoring of the generation and serial interval distributions (*32*). Symptomatic cases represent 86.5% of all SARS-CoV-2 infections in our data; among these patients, we observe longer generation intervals for cases isolated later in the course of their infection (Fig. 3A). The median generation interval increases from 4.0 days (IQR, 1.9 to 7.3 days) for cases isolated 2 day since symptom onset, to 7.0 days (IQR, 3.6 to 11.3 days) for those isolated more than 6 days after symptom onset (p<0.001, Mann-Whitney U test). We observe similar trends for the serial interval distribution (Fig. 3B). The median serial interval increases from 1.7 days (IQR, -1.6 to 4.8 days) for cases isolated less than 2 day after symptom onset, to 7.3 days (IQR, 3.4 to10.8 days) for those isolated more than 6 days after symptom onset (p<0.001, Mann-Whitney U test).

**Fig. 3.**
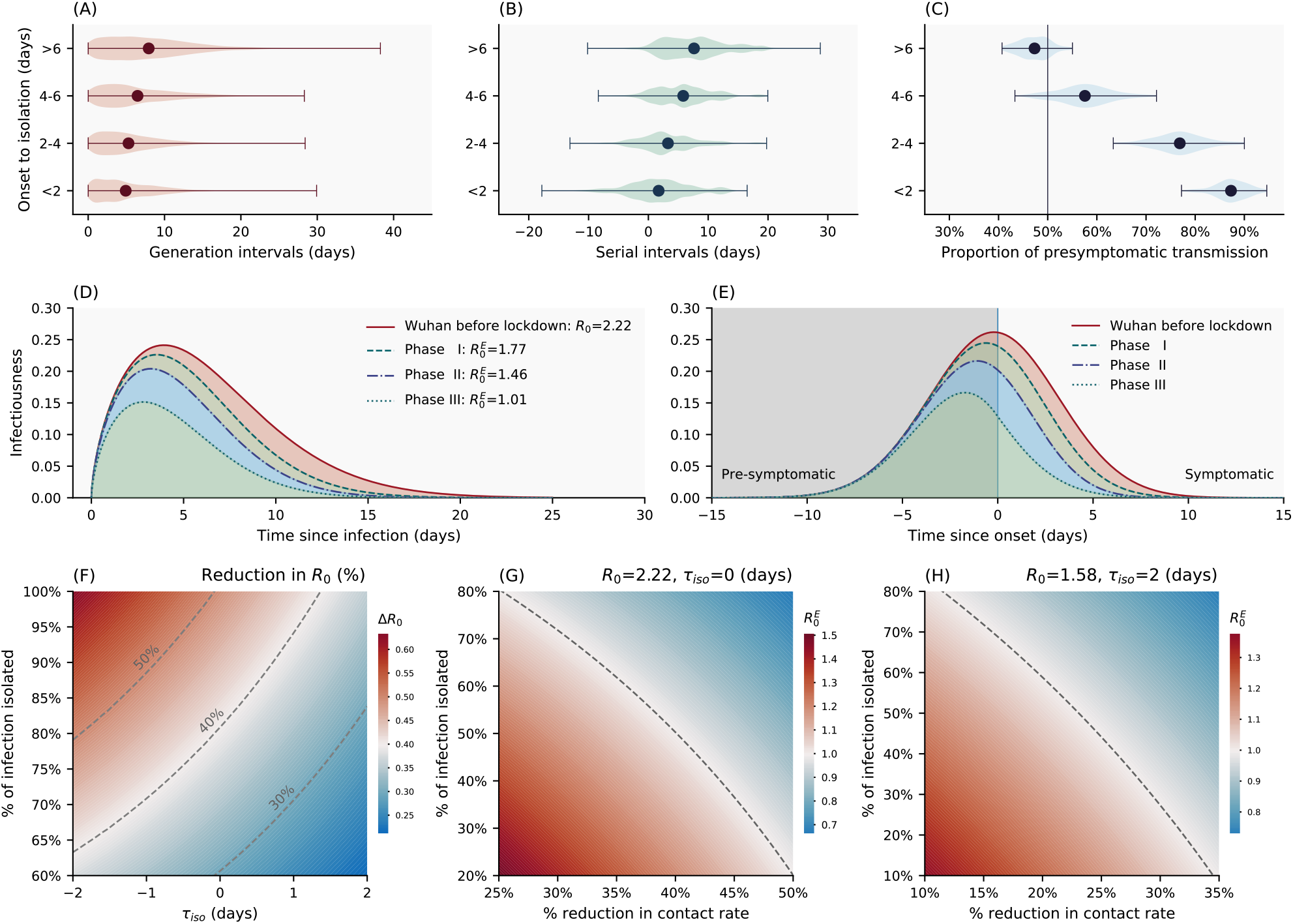
(A) Violin plot of the generation interval distributions stratified by time from symptom onset to isolation/pre-symptomatic quarantine, based on an ensemble of 100 realizations of the sampled transmission chains. (B) Same as A but for the serial interval distributions (C) Same as A but for the fraction of pre-symptomatic transmission, among all transmission events, with vertical line indicating 50% of pre-symptomatic transmission. Dots represent the mean and whiskers represents minimum and maximum. (D) Estimated average (over 100 realizations of sampled transmission chains) transmission risk of a SARS-CoV-2 infected individual since time of infection under four intervention scenarios: the red solid line represents an uncontrolled epidemic scenario modelled after the early epidemic dynamics in Wuhan before lockdown; the dashed lines represent scenarios where quarantine and case isolation are in place and mimic Phase I, II, and III of epidemic control in Hunan. The shapes of these curves match that of the generation interval distributions in each scenario while the areas under the curve are equal to the ratio of the baseline/effective basic reproduction numbers 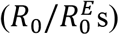. (E) Same as in D but with time since symptom onset on the x-axis (colors are as in (D)). The vertical line represents symptom onset. (F) Reduction (percentage) in the basic reproduction number as a function of mean time from symptom onset (or from peak infectiousness for asymptomatic cases) to isolation *τ_iso_* (x-axis) and fraction of SARS-CoV-2 infections being isolated (y-axis). The distribution of onset to isolation follows a normal distribution with mean *τ*_iso_ and standard deviation of 2 days. The dashed lines indicate 30%, 40% and 50% reductions in R0 under interventons. (G) Effective basic reproduction number as a function of population-level reduction in contact rates (i.e. through physical distancing, expressed as a percentage, x-axis) and isolation rate (fraction of total infections detected and further isolated). We assume baseline basic reproduction number *R*_0_ = 2.19, and a normal distribution for the distribution from onset to isolation with mean of 0 days and standard deviation of 2 days. The dashed line represents the epidemic threshold *R*^*E*^= 1. The blue area indicates region below the epidemic threshold (namely, controlled epidemic) and the red area indicates region above the epidemic threshold. (H) Same as in G but assuming *R*_0_ = 1.57 (a more optimistic estimate of R_0_ in Wuhan adjusted for reporting changes), and a normal distribution for the distribution from onset to isolation with mean of 2 days and standard deviation of 2 days.

Faster case isolation restricts transmission to the earlier stages of infection, thus inflating the contribution of pre-symptomatic transmission (Fig. 3C). The proportion of pre-symptomatic transmission is estimated at 87.3% (95% CI, 79.8% to 93.4%) if cases are isolated within 2 day of symptom onset, while this proportion decreases to 47.5% (95% CI, 41.4% to 53.3%) if cases are isolated more than 6 days after symptom onset (p<0.001, Mann-Whitney U test).

Next, we adjust for censoring due to case isolation and reconstruct the infectiousness profile of a typical SARS-CoV-2 patient in the absence of interventions. To do so, we characterize changes in the timeliness of case isolation over time in Hunan. Fig. S8 shows the distributions of time from symptom onset to isolation during three different phases of epidemic control, coinciding with major changes in COVID-19 case definition (*Phase I*: before Jan. 27^th^; *Phase II*: Jan. 27^th^ – Feb. 4^th^; *Phase III*: after Feb. 4^th^, Fig. S3) (*33*). In *Phase I*, 78% of cases were detected through passive surveillance; as a result, most cases were isolated after symptom onset (median time from onset to isolation 5.4 days, IQR (2.7, 8.2) days, Fig. S8A). In contrast, in *Phase III*, 66% of cases were detected through active contact tracing, shortening the median time from onset to isolation to -0.1 days (IQR (−2.9, 1.8) days, Fig. S8C). Timeliness of isolation is intermediate in *Phase II*. We use mathematical models (detailed in Materials and Methods) to dynamically adjust the serial interval distribution for censoring, and we apply the same approach to the time interval between a primary case’s symptom onset and onward transmission (Fig. S10). These censoring-adjusted distributions can be rescaled by the basic reproduction number *R*_0_ to reflect the risk of transmission of a typical SARS-CoV-2 case since the time of infection or since symptom onset (Fig 3D-E). We find that in the absence of interventions, infectiousness peaks near the time of symptoms onset (Fig. S10D). This is consistent with our regression analysis, where the higher risk of transmission is near symptom onset (Table 1).

### Evaluating the impact of individual and population-based interventions on SARS-CoV-2 transmission

Next, we use the estimated infectiousness profile of a typical SARS-CoV-2 infection (Fig. 3D-E) to evaluate the impact of case isolation on transmission. We first set a baseline reproduction number R_0_ for SARS-CoV-2 in the absence of control. Results from a recent study (*33*) suggest that the initial growth rate in Wuhan was 0.15 day^-1^ in raw case data (95% CI, 0.14 to 0.17), although the growth rate could be substantially lower (0.08 day^-1^) if changes in case definition are considered. Conservatively, we consider the upper value of the growth rate at 0.15 day^-1^ together with our generation interval distribution adjusted for censoring (Fig. S10C), to estimate R_0_. We obtain a baseline reproduction number *R*_0_ = 2.19 (95% CI, 2.08 to 2.36), using the renewal equation framework (*34*). This represents a typical scenario of unmitigated SARS-CoV-2 transmissibility in an urban setting. The reconstructed infectiousness profile in the absence of control is shown in solid red lines in Fig. 3D-E, with respect to time of infection and symptom onset respectively. Notably, we find that SARS-CoV-2 infectiousness peaks slightly before symptom onset (−0.1 days on average), with 87% of the overall infectiousness concentrated within ±5 days of symptom onset and 53% of the overall infectiousness in the pre-symptomatic phase (Fig. 3E).

Next, we evaluate the impact of case isolation on transmission by considering three different intervention scenarios mimicking the timeliness of isolation in the three phases of the Hunan epidemic control. We further assume that 100% of infections are detected and isolated and that isolation is fully protective (i.e., there is no onward transmission after the patient has been isolated/quarantined). The infectiousness profiles of the three intervention scenarios are shown in dashed lines in Fig. 3D-E. We find that the basic reproduction number decreases in all intervention scenarios, but the projected decrease is not sufficient to interrupt transmission (Fig. 3D, 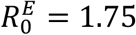 for *Phase I*, 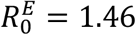 for *Phase II*, and 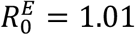 for *Phase III*).

We further relax the assumption of 100% case detection and isolation and relate changes in the basic reproduction number to two independent parameters measuring the strength of interventions: the effectiveness of case isolation and contact quarantine (measured as the fraction of total infections isolated) and the timeliness of isolation (measured as the delay from symptom onset to isolation, phase diagram in Fig. 3F). Dashed lines in Fig. 3F illustrate 30%, 40% and 50% of reduction in *R*_0_. To reduce *R*_0_ by half (the minimum amount of transmission reduction required to achieve control for a baseline *R*_0_∼2), 100% of infections would need to be isolated even if individuals are isolated as early as the day of symptom onset. In practice, epidemic control is unrealistic if case isolation and quarantine of close contacts are the only measures in place.

Our data support that case isolation and quarantine of close contacts are effective in reducing SARS-CoV-2 transmission, especially if these interventions occur early in the infection. To achieve epidemic control, however, these interventions need to be layered with additional population-level measures, including increased teleworking, reduced operation in the service industry, or broader adoption of face masks. The synergistic effects of these interventions are illustrated in Fig. 3G. We find that a 30% reduction in transmission from population-level measures would require a 70% case detection rate to achieve epidemic control, assuming that cases can be promptly isolated on average upon symptom presentation. Of note, a 30% reduction in transmission could also encompass the benefits of residual population-level immunity from the first wave of COVID-19, especially in hard-hit regions (*35, 36*). As a sensitivity analysis, we further consider a more optimistic scenario with a lower baseline *R*_0_ = 1.56, corresponding to an epidemic growth rate of 0.08 day^-1^ (95% CI, 0.06 to 0.10) in Wuhan (*33*), which is adjusted for reporting changes. As expected, control is much easier to achieve in this scenario: if detected SARS-CoV-2 infections are effectively isolated on average 2 days after symptom onset, a 25% population-level reduction in transmission coupled with a 42% infection isolation rate is sufficient to achieve control (Fig. 3H).

## Discussion

Detailed information on 1,178 SARS-CoV-2 infected individuals along with their 15,648 contacts has allowed us to dissect the behavioral and clinical drivers of SARS-CoV-2 transmission; to evaluate how transmission opportunities are modulated by individual and population-level interventions, and to characterize the typical infectiousness profile of a case. Informed by this understanding, particularly the importance of pre-symptomatic transmission, we have evaluated the plausibility of SARS-CoV-2 control through individual and population-based interventions.

Healthcare contacts pose the lowest risk of transmission in Hunan, suggesting that adequate protective measures against SARS-CoV-2 were taken in hospitals and medical observation centers (Table 1). The average risk of transmission scales positively with the closeness of social interactions: the average per-contact risk is lowest for community exposures (including contacts in the public transportation system and at food and entertainment venues), intermediate for social and extended family contacts, and highest in the household. The average transmission risk in the household is further elevated when intense physical distancing is enforced, and for contacts that last longer. These lines of evidence support that SARS-CoV-2 transmission is facilitated by close proximity, confined environment, and high frequency of contacts.

Regression analysis indicates a higher risk of transmission when an individual is exposed to a SARS-CoV-2 patient around the time of symptom onset, in line with our reconstructed infectiousness profile. These epidemiological findings are in agreement with viral shedding studies (*6, 37–40*). We estimate that overall in Hunan, 63% of all transmission events were from pre-symptomatic individuals, in concordance with other modeling studies (*6, 7, 10, 12, 41*). However, the estimated pre-symptomatic proportion is affected by case-based measures, including case isolation and contact quarantine. We estimate that the relative contribution of pre-symptomatic transmission drops to 52% in an uncontrolled scenario where case-based interventions are absent.

Case isolation reduces the “effective” infectious period of SARS-CoV-2 infected individuals by blocking contacts with susceptible individuals. We observe that faster isolation significantly reduces cumulative contact rates (CCRs) across contact types (Fig. 2E). We also observe shorter serial and generation intervals and a larger fraction of pre-symptomatic transmission when individuals are isolated faster (Fig. 3A-C). In contrast, population-level physical distancing measures have differential impacts on CCRs, decreasing CCRs for social and community contacts, while increasing CCRs in the household and family contacts. As a result, strict physical distancing confines the epidemic mostly to families and households (see also Fig. S7). The precise impact of physical distancing on transmission is difficult to separate from that of individual-based interventions. However, our analysis suggests that physical distancing changes the topology of the transmission network by affecting the number and duration of interactions. Interestingly, the topology of the household contact network is highly clustered (*42*), and theoretical studies have shown that high clustering hinders epidemic spread (*43, 44*). These higher-order topological changes could contribute to reducing transmission beyond the effects expected from an overall reduction in CCRs. In parallel, the effectiveness of physical distancing measures on reducing COVID-19 transmission has been demonstrated in empirical data from China (*16, 45*) and elsewhere (*46*).

We have explored the feasibility of SARS-CoV-2 epidemic control against two important metrics related to case isolation and contact quarantine: the timeliness of isolation and the infection detection rate (Fig. 3F). For a baseline transmission scenario compatible with the initial growth phase of the epidemic in Wuhan, we find that epidemic control solely relying on isolation and quarantine is difficult to achieve. Layering these interventions with moderate physical distancing makes control more likely over a range of plausible parameters - a situation that could be further improved by residual immunity from the first wave of SARS-CoV-2 activity (*35, 36*). Successful implementation of contact tracing requires a low-level of active infections in the community, as the number of contacts to be monitored is several folds the number of infections (∼13 contacts were traced for each SARS-CoV-2 infected individual in Hunan). The timing of easing of lockdown measures should align with the capacities of testing and contact tracing efforts, relative to the number of active infections in the community. In parallel, technology-based approaches can also facilitate these efforts (*7, 47*).

Overall, we find that case isolation and quarantine successfully blocked transmission to close contacts in Hunan, with an estimated 4.3% of transmission occurring after SARS-CoV-2 patients were isolated. In this setting, all SARS-CoV-2 infections were managed under medical isolation in dedicated hospitals regardless of clinical severity, while contacts were quarantined in designated medical observation centers. Self-regulated isolation and quarantine at home, however, may not be as effective and a higher proportion of onward transmission should be expected.

Several caveats are worth noting. We could not evaluate the risk of transmission in schools, workplaces, conferences, prisons, or factories, as no contacts in these settings were reported in the Hunan dataset. Our study is likely underpowered to assess the transmission potential of asymptomatic individuals given the relatively small fraction of these infections in our data (13.5% overall and 22.1% of infections captured through contact tracing). There is no statistical support for decreased transmission from asymptomatic individuals (Fig. S3A), although we observe a positive, but non-significant gradient in average transmission risk with disease severity. Evidence from viral shedding studies is conflicting; viral load appears independent of clinical severity in some studies (*6, 23, 38, 48*) while others suggest faster viral clearance in asymptomatic individuals (*49*).

Another limitation relates to changes in testing practices for contacts of primary cases. Testing was initially limited to contacts exhibiting symptoms, and this condition was relaxed after February 7^th^. The early testing scheme may lead to underestimation of susceptibility in children, as younger individuals are less likely to develop SARS-CoV-2 symptoms (*50*). However, reassuringly, sensitivity analyses indicate that the age gradient of susceptibility is preserved even after stratification for changes in testing protocol, and our estimate of a lower susceptibility to infection of children (individuals aged 0-12 years) as compared to adults is stable in the period with broadened testing (Fig. S4). Overall, the contribution of asymptomatic infections to transmission remains debated but has profound implications on the feasibility of control through individual-based interventions. Careful serological studies combined with virologic testing in households and other controlled environments are needed to fully resolve the role of asymptomatic infections and viral shedding on transmission.

In conclusion, detailed contact tracing data illuminate important heterogeneities in SARS-CoV-2 transmission driven by biology and behavior, modulated by the impact of interventions. Crucially, and in contrast to SARS-CoV-1, the ability of SARS-CoV-2 to transmit during the host’s pre-symptomatic phase makes it particularly difficult to achieve epidemic control (*51*). Our risk factor estimates can provide useful evidence to guide the design of more targeted and sustainable mitigation strategies, while our reconstructed transmission kinetics will help calibrate further modeling efforts.

## Materials and Methods Summary

We combined individual-level data on 1,178 SARS-CoV-2 infections with detailed diaries of exposures collected through contact tracing efforts in Hunan, China, to stochastically reconstruct transmission chains and infer infection times. Reconstructed transmission chains had to be compatible with highly resolved individual level data on symptom onset dates, daily records of exposure to infected contacts, and travel history to high risk regions. Based on the reconstructed transmission chains, we characterize the distribution of key SARS-CoV-2 transmission parameters, including the number of secondary cases, the generation and serial intervals, and the interval from infection or symptom onset to isolation, at different stages of the epidemic. To further understand the drivers of transmission heterogeneity and the dispersion in the number of secondary cases, we study the degree distribution of SARS-CoV2 infected individuals, the duration of exposures, and the age specific contact patterns between infectors and infectees, separately by contact type (household, family, community, transportation, healthcare). We also use logistic regression analysis to model the per-contact risk of transmission, with contact type and duration, symptoms, demographic factors, and different periods of the outbreak as covariates. Missing data are addressed through multivariate imputation algorithms. We conduct sensitivity analyses to test the robustness of regression results.

We next use our data to model the synergistic effects of case-based and population-level interventions on transmission. We reconstruct the average infectiousness profile of a SARS-CoV-2 infection, after adjusting for the truncation effects of case isolation. Based on the estimated infectiousness profile, we use mathematical models to estimate the effect of layered interventions on transmission (measured as changes in the effective reproduction number). We consider different intensities of population-level physical distancing, case detection, and timeliness of isolation/quarantine. A full description of the Materials and Methods is provided in the Supplementary Material.

## Data Availability

The original database containing confidential patient information cannot be made public however, we created a synthetic mock database to demonstrate the underlying data structure. All code, mock database, along with de-identified data to reproduce all figures in the main text and supplementary material are publicly available at doi:10.5281/ZENODO.4129864

https://doi.org/10.5281/zenodo.4129863

## Acknowledgements

The authors acknowledge Dr Christophe Fraser from the University of Oxford, Dr David Spiro from Fogarty International Center, National Institutes of Health, and Dr Peter Kilmarx from Fogarty International Center, National Institutes of Health for their helpful comments on the manuscript. This article does not necessarily represent the views of the NIH or the U.S. government.

## Author contributions

C.V. and H.Y. are joint senior authors. K.S., C.V. and H.Y. designed the experiments. L.G., W.W, and Y.W. collected data. K.S., W.W., L.G., and Y.W. analyzed the data. K.S., W.W., L.G., Y.W., K.L., L.R., Z.Z., X.C., S.Z., Y.H., Q.S., Z.L., M.L., A.V., M.A., C.V., and H.Y. interpreted the results. K.S., W.W., L.G., Y.W., M.L., A.V., M.A., C.V. and H.Y. wrote the manuscript.

## Ethics statement

The collection of specimens, epidemiological and clinical data for SARS-CoV-2 infected individuals was part of a continuing public health investigation of an emerging outbreak, defined by the *Protocol on the Prevention and Control of COVID-19* established by the National Health Commission of the People’s Republic of China (*52*). Thus, collection of epidemiological and clinical data was exempt from institutional review board assessment. This study was approved by the Institutional Review Board of Hunan Provincial Center for Disease Control and Prevention (IRB#2020005). Data were deidentified, and informed consent was waived with completion of an appropriate institutional form.

## Data and materials availability

The original database containing confidential patient information cannot be made public; however, we have created a synthetic database to demonstrate the structure of the underlying data. All code, the mock database, along with de-identified data to reproduce all figures in the main text and supplementary material are publicly available at (*53*). This work is licensed under a Creative Commons Attribution 4.0 International (CC BY 4.0) license, which permits unrestricted use, distribution, and reproduction in any medium, provided the original work is properly cited. To view a copy of this license, visit https://creativecommons.org/licenses/by/4.0/. This license does not apply to figures/photos/artwork or other content included in the article that is credited to a third party; obtain authorization from the rights holder before using such material.

## Supplementary Materials

Materials and Methods

Figures S1-S11 Tables S1-S5

References (*12, 25, 33, 34, 51,52, 54-61*)

## References and Notes

1. W. Guan, Z. Ni, Y. Hu, W. Liang, C. Ou, J. He, L. Liu, H. Shan, C. Lei, D. S. C. Hui, B. Du, L. Li, G. Zeng, K.-Y. Yuen, R. Chen, C. Tang, T. Wang, P. Chen, J. Xiang, S. Li, J. Wang, Z. Liang, Y. Peng, L. Wei, Y. Liu, Y. Hu, P. Peng, J. Wang, J. Liu, Z. Chen, G. Li, Z. Zheng, S. Qiu, J. Luo, C. Ye, S. Zhu, N. Zhong, Clinical Characteristics of Coronavirus Disease 2019 in China. N. Engl. J. Med. 382, 1708–1720 (2020).

2. J. T. Wu, K. Leung, M. Bushman, N. Kishore, R. Niehus, P. M. de Salazar, B. J. Cowling, M. Lipsitch, G. M. Leung, Estimating clinical severity of COVID-19 from the transmission dynamics in Wuhan, China. Nat. Med. 26, 506–510 (2020).

3. G. Grasselli, A. Zangrillo, A. Zanella, M. Antonelli, L. Cabrini, A. Castelli, D. Cereda, A. Coluccello, G. Foti, R. Fumagalli, G. Iotti, N. Latronico, L. Lorini, S. Merler, G. Natalini, Piatti, M. V. Ranieri, A. M. Scandroglio, E. Storti, M. Cecconi, A. Pesenti, Baseline Characteristics and Outcomes of 1591 Patients Infected with SARS-CoV-2 Admitted to ICUs of the Lombardy Region, Italy. JAMA - J. Am. Med. Assoc. 323, 1574–1581 (2020).

4. R. Verity, L. C. Okell, I. Dorigatti, P. Winskill, C. Whittaker, N. Imai, G. Cuomo-Dannenburg, H. Thompson, P. G. T. Walker, H. Fu, A. Dighe, J. T. Griffin, M. Baguelin, S. Bhatia, A. Boonyasiri, A. Cori, Z. Cucunubá, R. FitzJohn, K. Gaythorpe, W. Green, A. Hamlet, W. Hinsley, D. Laydon, G. Nedjati-Gilani, S. Riley, S. van Elsland, E. Volz, H. Wang, Y. Wang, X. Xi, C. A. Donnelly, A. C. Ghani, N. M. Ferguson, Estimates of the severity of coronavirus disease 2019: a model-based analysis. Lancet Infect. Dis. 20, 669– 677 (2020).

5. K. Sun, J. Chen, C. Viboud, Early epidemiological analysis of the coronavirus disease 2019 outbreak based on crowdsourced data: a population-level observational study. Lancet Digit. Heal. 2, e201–e208 (2020).

6. X. He, E. H. Y. Lau, P. Wu, X. Deng, J. Wang, X. Hao, Y. C. Lau, J. Y. Wong, Y. Guan, X. Tan, X. Mo, Y. Chen, B. Liao, W. Chen, F. Hu, Q. Zhang, M. Zhong, Y. Wu, L. Zhao, F. Zhang, B. J. Cowling, F. Li, G. M. Leung, Temporal dynamics in viral shedding and transmissibility of COVID-19. Nat. Med. 26, 672–675 (2020).

7. L. Ferretti, C. Wymant, M. Kendall, L. Zhao, A. Nurtay, L. Abeler-Dörner, M. Parker, D. Bonsall, C. Fraser, Quantifying SARS-CoV-2 transmission suggests epidemic control with digital contact tracing. Science (80-.). 368 (2020), doi:10.1126/science.abb6936.

8. A. Kimball, K. M. Hatfield, M. Arons, A. James, J. Taylor, K. Spicer, A. C. Bardossy, L. P. Oakley, S. Tanwar, Z. Chisty, J. M. Bell, M. Methner, J. Harney, J. R. Jacobs, C. M. Carlson, H. P. McLaughlin, N. Stone, S. Clark, C. Brostrom-Smith, L. C. Page, M. Kay, J. Lewis, D. Russell, B. Hiatt, J. Gant, J. S. Duchin, T. A. Clark, M. A. Honein, S. C. Reddy, J. A. Jernigan, A. Baer, L. M. Barnard, E. Benoliel, M. S. Fagalde, J. Ferro, H. G. Smith, E. Gonzales, N. Hatley, G. Hatt, M. Hope, M. Huntington-Frazier, V. Kawakami, J. L. Lenahan, M. D. Lukoff, E. B. Maier, S. McKeirnan, P. Montgomery, J. L. Morgan, L. A. Mummert, S. Pogosjans, F. X. Riedo, L. Schwarcz, D. Smith, S. Stearns, K. J. Sykes, H. Whitney, H. Ali, M. Banks, A. Balajee, E. J. Chow, B. Cooper, D. W. Currie, J. Dyal, J. Healy, M. Hughes, T. M. McMichael, L. Nolen, C. Olson, A. K. Rao, K. Schmit, N. G. Schwartz, F. Tobolowsky, R. Zacks, S. Zane, Asymptomatic and presymptomatic SARS-COV-2 infections in residents of a long-term care skilled nursing facility - King County, Washington, March 2020. Morb. Mortal. Wkly. Rep. 69, 377–381 (2020).

9. W. E. Wei, Z. Li, C. J. Chiew, S. E. Yong, M. P. Toh, V. J. Lee, Presymptomatic transmission of SARS-CoV-2 — Singapore, January 23–March 16, 2020. Morb. Mortal. Wkly. Rep. 69, 411–415 (2020).

10. Z. Du, X. Xu, Y. Wu, L. Wang, B. J. Cowling, L. A. Meyers, Serial Interval of COVID-19 among Publicly Reported Confirmed Cases. Emerg. Infect. Dis. 26, 1341–1343 (2020).

11. E. Lavezzo, E. Franchin, C. Ciavarella, G. Cuomo-Dannenburg, L. Barzon, C. Del Vecchio, L. Rossi, R. Manganelli, A. Loregian, N. Navarin, D. Abate, M. Sciro, S. Merigliano, E. Decanale, M. C. Vanuzzo, F. Saluzzo, F. Onelia, M. Pacenti, S. Parisi, G. Carretta, D. Donato, L. Flor, S. Cocchio, G. Masi, A. Sperduti, L. Cattarino, R. Salvador, K. A. M. Gaythorpe, I. C. L. C.-19 R. Team, A. R. Brazzale, S. Toppo, M. Trevisan, V. Baldo, C. A. Donnelly, N. M. Ferguson, I. Dorigatti, A. Crisanti, Suppression of COVID-19 outbreak in the municipality of Vo, Italy. medRxiv (2020), doi:10.1101/2020.04.17.20053157.

12. S. Hu, W. Wang, Y. Wang, M. Litvinova, K. Luo, L. Ren, Q. Sun, X. Chen, G. Zeng, J. Li, L. Liang, Z. Deng, W. Zheng, M. Li, H. Yang, J. Guo, K. Wang, X. Chen, Z. Liu, H. Yan, H. Shi, Z. Chen, Y. Zhou, K. Sun, A. Vespignani, C. Viboud, L. Gao, M. Ajelli, H. Yu, Infectivity, susceptibility, and risk factors associated with SARS-CoV-2 transmission under intensive contact tracing in Hunan, China. medRxiv (2020), doi:10.1101/2020.07.23.20160317.

13. Q. Bi, Y. Wu, S. Mei, C. Ye, X. Zou, Z. Zhang, X. Liu, L. Wei, S. A. Truelove, T. Zhang, W. Gao, C. Cheng, X. Tang, X. Wu, Y. Wu, B. Sun, S. Huang, Y. Sun, J. Zhang, T. Ma, J. Lessler, T. Feng, Epidemiology and transmission of COVID-19 in 391 cases and 1286 of their close contacts in Shenzhen, China: a retrospective cohort study. Lancet Infect. Dis. (2020), doi:10.1016/S1473-3099(20)30287-5.

14. A. Pan, L. Liu, C. Wang, H. Guo, X. Hao, Q. Wang, J. Huang, N. He, H. Yu, X. Lin, S. Wei, T. Wu, Association of Public Health Interventions With the Epidemiology of the COVID-19 Outbreak in Wuhan, China. JAMA. 323, 1915 (2020).

15. B. J. Cowling, S. T. Ali, T. W. Y. Ng, T. K. Tsang, J. C. M. Li, M. W. Fong, Q. Liao, M. Y. Kwan, S. L. Lee, S. S. Chiu, J. T. Wu, P. Wu, G. M. Leung, Impact assessment of non-pharmaceutical interventions against coronavirus disease 2019 and influenza in Hong Kong: an observational study. Lancet Public Heal. 5, e279–e288 (2020).

16. J. Zhang, M. Litvinova, Y. Liang, Y. Wang, W. Wang, S. Zhao, Q. Wu, S. Merler, C. Viboud, A. Vespignani, M. Ajelli, H. Yu, Changes in contact patterns shape the dynamics of the COVID-19 outbreak in China. Science (80-.). 368, 1481–1486 (2020).

17. S. W. Park, K. Sun, C. Viboud, B. T. Grenfell, J. Dushoff, Potential roles of social distancing in mitigating the spread of coronavirus disease 2019 (COVID-19) in South Korea. medRxiv (2020), doi:10.1101/2020.03.27.20045815.

18. A. Aleta, D. Martin-Corral, A. P. y Piontti, M. Ajelli, M. Litvinova, M. Chinazzi, N. E. Dean, M. E. Halloran, I. M. Longini, S. Merler, A. Pentland, A. Vespignani, E. Moro, Y. Moreno, D. Mart, A. Pastore, M. Ajelli, M. Litvinova, M. Chinazzi, N. E. Dean, M. E. Halloran, Modeling the impact of social distancing testing contact tracing and household quarantine on second-wave scenarios of the COVID-19 epidemic. medRxiv (2020), doi:10.1101/2020.05.06.20092841.

19. J. Zhang, M. Litvinova, W. Wang, Y. Wang, X. Deng, X. Chen, M. Li, W. Zheng, L. Yi, X. Chen, Q. Wu, Y. Liang, X. Wang, J. Yang, K. Sun, I. M. Longini, M. E. Halloran, P. Wu, B. J. Cowling, S. Merler, C. Viboud, A. Vespignani, M. Ajelli, H. Yu, Evolving epidemiology and transmission dynamics of coronavirus disease 2019 outside Hubei province, China: a descriptive and modelling study. Lancet Infect. Dis. 20, 793–802 (2020).

20. K. Leung, J. T. Wu, D. Liu, G. M. Leung, First-wave COVID-19 transmissibility and severity in China outside Hubei after control measures, and second-wave scenario planning: a modelling impact assessment. Lancet. 395, 1382–1393 (2020).

21. M. Chinazzi, J. T. Davis, M. Ajelli, C. Gioannini, M. Litvinova, S. Merler, A. Pastore y Piontti, K. Mu, L. Rossi, K. Sun, C. Viboud, X. Xiong, H. Yu, M. Elizabeth Halloran, I. M. Longini, A. Vespignani, The effect of travel restrictions on the spread of the 2019 novel coronavirus (COVID-19) outbreak. Science (80-.). 368, 395–400 (2020).

22. H. Tian, Y. Liu, Y. Li, C. H. Wu, B. Chen, M. U. G. Kraemer, B. Li, J. Cai, B. Xu, Q. Yang, B. Wang, P. Yang, Y. Cui, Y. Song, P. Zheng, Q. Wang, O. N. Bjornstad, R. Yang, B. T. Grenfell, O. G. Pybus, C. Dye, An investigation of transmission control measures during the first 50 days of the COVID-19 epidemic in China. Science (80-.). 368, 638– 642 (2020).

23. D. Cereda, M. Tirani, F. Rovida, V. Demicheli, M. Ajelli, P. Poletti, F. Trentini, G. Guzzetta, V. Marziano, A. Barone, M. Magoni, S. Deandrea, G. Diurno, M. Lombardo, M. Faccini, A. Pan, R. Bruno, E. Pariani, G. Grasselli, A. Piatti, M. Gramegna, F. Baldanti, A. Melegaro, S. Merler, The early phase of the COVID-19 outbreak in Lombardy, Italy. arXiv (2020) (available at http://arxiv.org/abs/2003.09320).

24. A. J. Kucharski, T. W. Russell, C. Diamond, Y. Liu, J. Edmunds, S. Funk, R. M. Eggo, F. Sun, M. Jit, J. D. Munday, N. Davies, A. Gimma, K. van Zandvoort, H. Gibbs, J. Hellewell, C. I. Jarvis, S. Clifford, B. J. Quilty, N. I. Bosse, S. Abbott, P. Klepac, S. Flasche, Early dynamics of transmission and control of COVID-19: a mathematical modelling study. Lancet Infect. Dis. 20, 553–558 (2020).

25. Baidu.com, Baidu Qianxi: Within city mobility index. https://qianxi.baidu.com/2020/ (2020).

26. E. Goldstein, M. Lipsitch, medRxiv, in press, doi:10.1101/2020.07.19.20157362.

27. R. M. Viner, O. T. Mytton, C. Bonell, G. J. Melendez-Torres, J. Ward, L. Hudson, C. Waddington,J. Thomas, S. Russell, F. van der Klis, A. Koirala, S. Ladhani, J. Panovska-Griffiths, N. G. Davies, R. Booy, R. M. Eggo, Susceptibility to SARS-CoV-2 Infection Among Children and Adolescents Compared With Adults. JAMA Pediatr. (2020), doi:10.1001/jamapediatrics.2020.4573.

28. J. Mossong, N. Hens, M. Jit, P. Beutels, K. Auranen, R. Mikolajczyk, M. Massari, S. Salmaso, G. S. Tomba, J. Wallinga, J. Heijne, M. Sadkowska-Todys, M. Rosinska, W. J. Edmunds, Social Contacts and Mixing Patterns Relevant to the Spread of Infectious Diseases. PLoS Med. 5, e74 (2008).

29. M. Litvinova, Q. H. Liu, E. S. Kulikov, M. Ajelli, Reactive school closure weakens the network of social interactions and reduces the spread of influenza. Proc. Natl. Acad. Sci. U. S. A. 116, 13174–13181 (2019).

30. Q. Li, X. Guan, P. Wu, X. Wang, L. Zhou, Y. Tong, R. Ren, K. S. M. Leung, E. H. Y. Lau, J. Y. Wong, X. Xing, N. Xiang, Y. Wu, C. Li, Q. Chen, D. Li, T. Liu, J. Zhao, M. Liu, W. Tu, C. Chen, L. Jin, R. Yang, Q. Wang, S. Zhou, R. Wang, H. Liu, Y. Luo, Y. Liu, G. Shao, H. Li, Z. Tao, Y. Yang, Z. Deng, B. Liu, Z. Ma, Y. Zhang, G. Shi, T. T. Y. Lam, J. T. Wu, G. F. Gao, B. J. Cowling, B. Yang, G. M. Leung, Z. Feng, Early Transmission Dynamics in Wuhan, China, of Novel Coronavirus–Infected Pneumonia. N. Engl. J. Med. 382, 1199–1207 (2020).

31. T. Ganyani, C. Kremer, D. Chen, A. Torneri, C. Faes, J. Wallinga, N. Hens, Estimating the generation interval for coronavirus disease (COVID-19) based on symptom onset data, March 2020. Eurosurveillance. 25, 2000257 (2020).

32. S. T. Ali, L. Wang, E. H. Y. Lau, X.-K. Xu, Z. Du, Y. Wu, G. M. Leung, B. J. Cowling, Serial interval of SARS-CoV-2 was shortened over time by nonpharmaceutical interventions. Science (2020), doi:10.1126/science.abc9004.

33. T. K. Tsang, P. Wu, Y. Lin, E. H. Y. Lau, G. M. Leung, B. J. Cowling, Effect of changing case definitions for COVID-19 on the epidemic curve and transmission parameters in mainland China: a modelling study. Lancet Public Heal. 5, e289–e296 (2020).

34. J. Wallinga, M. Lipsitch, How generation intervals shape the relationship between growth rates and reproductive numbers. Proc. R. Soc. B Biol. Sci. 274, 599–604 (2007).

35. E. S. Rosenberg, J. M. Tesoriero, E. M. Rosenthal, R. Chung, M. A. Barranco, L. M. Styer, M. M. Parker, S.-Y. John Leung, J. E. Morne, D. Greene, D. R. Holtgrave, D. Hoefer, J. Kumar, T. Udo, B. Hutton, H. A. Zucker, Cumulative incidence and diagnosis of SARS-CoV-2 infection in New York. Ann. Epidemiol. (2020), doi:10.1016/j.annepidem.2020.06.004.

36. S. Stringhini, A. Wisniak, G. Piumatti, A. S. Azman, S. A. Lauer, H. Baysson, D. De Ridder, D. Petrovic, S. Schrempft, K. Marcus, S. Yerly, I. Arm Vernez, O. Keiser, S. Hurst, K. M. Posfay-Barbe, D. Trono, D. Pittet, L. Gétaz, F. Chappuis, I. Eckerle, N. Vuilleumier, B. Meyer, A. Flahault, L. Kaiser, I. Guessous, Seroprevalence of anti-SARS-CoV-2 IgG antibodies in Geneva, Switzerland (SEROCoV-POP): a population-based study. Lancet (2020), doi:10.1016/s0140-6736(20)31304-0.

37. R. Wölfel, V. M. Corman, W. Guggemos, M. Seilmaier, S. Zange, M. A. Müller, D. Niemeyer, T. C. Jones, P. Vollmar, C. Rothe, M. Hoelscher, T. Bleicker, S. Brünink, J. Schneider, R. Ehmann, K. Zwirglmaier, C. Drosten, C. Wendtner, Virological assessment of hospitalized patients with COVID-2019. Nature. 581, 465–469 (2020).

38. K. K. W. To, O. T. Y. Tsang, W. S. Leung, A. R. Tam, T. C. Wu, D. C. Lung, C. C. Y. Yip, J. P. Cai, J. M. C. Chan, T. S. H. Chik, D. P. L. Lau, C. Y. C. Choi, L. L. Chen, W. M. Chan, K. H. Chan, J. D. Ip, A. C. K. Ng, R. W. S. Poon, C. T. Luo, V. C. C. Cheng, J. F. W. Chan, I. F. N. Hung, Z. Chen, H. Chen, K. Y. Yuen, Temporal profiles of viral load in posterior oropharyngeal saliva samples and serum antibody responses during infection by SARS-CoV-2: an observational cohort study. Lancet Infect. Dis. 20, 565–574 (2020).

39. J. J. A. Van Kampen, D. A. M. C. Van De Vijver, P. L. A. Fraaij, B. L. Haagmans, M. M. Lamers, M. Sc, N. Okba, J. P. C. Van Den Akker, H. Endeman, D. A. M. P. J. Gommers, J. J. Cornelissen, R. A. S. Hoek, M. M. Van Der Eerden, D. A. Hesselink, H. J. Metselaar, A. Verbon, J. E. M. De Steenwinkel, G. I. Aron, E. C. M. Van Gorp, S. Van Boheemen, J. C. Voermans, C. A. B. Boucher, R. Molenkamp, M. P. G. Koopmans, C. Geurtsvankessel, A. A. Van Der Eijk, medRxiv, in press, doi:10.1101/2020.06.08.20125310.

40. A. Singanayagam, M. Patel, A. Charlett, J. Lopez Bernal, V. Saliba, J. Ellis, S. Ladhani, M. Zambon, R. Gopal, Duration of infectiousness and correlation with RT-PCR cycle threshold values in cases of COVID-19, England, January to May 2020. Eurosurveillance. 25, 2001483 (2020).

41. Y. Liu, S. Funk, S. Flasche, The contribution of pre-symptomatic infection to the transmission dynamics of COVID-2019. Wellcome Open Res. (2020), doi:10.12688/wellcomeopenres.15788.1.

42. Q. H. Liu, M. Ajelli, A. Aleta, S. Merler, Y. Moreno, A. Vespignani, Measurability of the epidemic reproduction number in data-driven contact networks. Proc. Natl. Acad. Sci. U. S. A. 115, 12680–12685 (2018).

43. R. Pastor-Satorras, C. Castellano, P. Van Mieghem, A. Vespignani, Epidemic processes in complex networks. Rev. Mod. Phys. 87, 925 (2015).

44. E. M. Volz, J. C. Miller, A. Galvani, L. Meyers, Effects of heterogeneous and clustered contact patterns on infectious disease dynamics. PLoS Comput. Biol. 7, e1002042 (2011).

45. N. M. Ferguson, K. E. C. Ainslie, C. E. Walters, H. Fu, S. Bhatia, H. Wang, X. Xi, M. Baguelin, S. Bhatt, A. Boonyasiri, O. Boyd, L. Cattarino, C. Ciavarella, Z. Cucunuba, G. Cuomo-Dannenburg, A. Dighe, I. Dorigatti, S. L. van Elsland, R. FitzJohn, K. Gaythorpe, C. Ghani, W. Green, A. Hamlet, W. Hinsley, N. Imai, D. Jorgensen, E. Knock, D. Laydon, G. Nedjati-Gilani, L. C. Okell, I. Siveroni, H. A. Thompson, H. J. T. Unwin, R. Verity, M. Vollmer, P. G. T. Walker, Y. Wang, O. J. Watson, C. Whittaker, P. Winskill, C. A. Donnelly, S. Riley, Evidence of initial success for China exiting COVID-19 social distancing policy after achieving containment. Wellcome Open Res. 5, 81 (2020).

46. S. Flaxman, S. Mishra, A. Gandy, H. J. T. Unwin, T. A. Mellan, H. Coupland, C. Whittaker, H. Zhu, T. Berah, J. W. Eaton, M. Monod, P. N. Perez-Guzman, N. Schmit, L. Cilloni, K. E. C. Ainslie, M. Baguelin, A. Boonyasiri, O. Boyd, L. Cattarino, L. V. Cooper, Z. Cucunubá, G. Cuomo-Dannenburg, A. Dighe, B. Djaafara, I. Dorigatti, S. L. van Elsland, R. G. FitzJohn, K. A. M. Gaythorpe, L. Geidelberg, N. C. Grassly, W. D. Green, T. Hallett, A. Hamlet, W. Hinsley, B. Jeffrey, E. Knock, D. J. Laydon, G. Nedjati-Gilani, P. Nouvellet, K. V. Parag, I. Siveroni, H. A. Thompson, R. Verity, E. Volz, C. E. Walters, H. Wang, Y. Wang, O. J. Watson, P. Winskill, X. Xi, P. G. Walker, A. C. Ghani, C. A. Donnelly, S. M. Riley, M. A. C. Vollmer, N. M. Ferguson, L. C. Okell, S. Bhatt, Estimating the effects of non-pharmaceutical interventions on COVID-19 in Europe. Nature (2020), doi:10.1038/s41586-020-2405-7.

47. J. Hellewell, S. Abbott, A. Gimma, N. I. Bosse, C. I. Jarvis, T. W. Russell, J. D. Munday, J. Kucharski, W. J. Edmunds, F. Sun, S. Flasche, B. J. Quilty, N. Davies, Y. Liu, S. Clifford, P. Klepac, M. Jit, C. Diamond, H. Gibbs, K. van Zandvoort, S. Funk, R. M. Eggo, Feasibility of controlling COVID-19 outbreaks by isolation of cases and contacts. Lancet Glob. Heal. 8, e488–e496 (2020).

48. Q.-X. Long, X.-J. Tang, Q.-L. Shi, Q. Li, H.-J. Deng, J. Yuan, J.-L. Hu, W. Xu, Y. Zhang, F.-J. Lv, K. Su, F. Zhang, J. Gong, B. Wu, X.-M. Liu, J.-J. Li, J.-F. Qiu, J. Chen, A.-L. Huang, Clinical and immunological assessment of asymptomatic SARS-CoV-2 infections. Nat. Med. (2020), doi:10.1038/s41591-020-0965-6.

49. N. V. V. Chau, V. T. Lam, N. T. Dung, L. M. Yen, N. N. Q. Minh, L. M. Hung, N. M. Ngoc, N. T. Dung, D. N. H. Man, L. A. Nguyet, L. T. H. Nhat, L. N. T. Nhu, N. T. H. Ny, N. T. T. Hong, E. Kestelyn, N. T. P. Dung, N. T. Phong, T. C. Xuan, T. T. Hien, T. N. H. Tu, R. B. Geskus, T. T. Thanh, N. T. Truong, N. T. Binh, T. C. Thuong, G. Thwaites, L. Van Tan, The natural history and transmission potential of asymptomatic SARS-CoV-2 infection. medRxiv (2020), doi:10.1101/2020.04.27.20082347.

50. P. Poletti, M. Tirani, D. Cereda, F. Trentini, G. Guzzetta, G. Sabatino, V. Marziano, A. Castrofino, F. Grosso, G. Del Castillo, R. Piccarreta, A. L. C.-19 T. Force, A. Andreassi, Melegaro, M. Gramegna, M. Ajelli, S. Merler, Probability of symptoms and critical disease after SARS-CoV-2 infection. arXiv (2020) (available at http://arxiv.org/abs/2006.08471).

51. C. Fraser, S. Riley, R. M. Anderson, N. M. Ferguson, Factors that make an infectious disease outbreak controllable. Proc. Natl. Acad. Sci. U. S. A. 101, 6146–6151 (2004).

52. Protocol on Prevention and Control of COVID-19 (Edition 6). Natl. Heal. Comm. People’s Repub. China (2020), (available at http://www.nhc.gov.cn/jkj/s3577/202003/4856d5b0458141fa9f376853224d41d7.shtml).

53. K. Sun, W. Wang, L. Gao, Y. Wang, K. Luo, L. Ren, Z. Zhang, X. Chen, S. Zhao, Y. Huang, Q. Sun, Z. Liu, M. Litvinova, A. Vespignani, M. Ajelli, C. Viboud, H. Yu, Code and data sharing for manuscript titled “Transmission heterogeneities, kinetics, and controllability of SARS-CoV-2” (2020),, doi:10.5281/ZENODO.4129864.

54. B. Nikolay, H. Salje, M. J. Hossain, A. K. M. D. Khan, H. M. S. Sazzad, M. Rahman, P. Daszak, U. Ströher, J. R. C. Pulliam, A. M. Kilpatrick, S. T. Nichol, J. D. Klena, S. Sultana, S. Afroj, S. P. Luby, S. Cauchemez, E. S. Gurley, Transmission of Nipah Virus — 14 Years of Investigations in Bangladesh. N. Engl. J. Med. 380, 1804–1814 (2019).

55. J. Agua-Agum, A. Ariyarajah, B. Aylward, L. Bawo, P. Bilivogui, I. M. Blake, R. J. Brennan, A. Cawthorne, E. Cleary, P. Clement, R. Conteh, A. Cori, F. Dafae, B. Dahl, J.-M. Dangou, B. Diallo, C. A. Donnelly, I. Dorigatti, C. Dye, T. Eckmanns, M. Fallah, N. M. Ferguson, L. Fiebig, C. Fraser, T. Garske, L. Gonzalez, E. Hamblion, N. Hamid, S. Hersey, W. Hinsley, A. Jambei, T. Jombart, D. Kargbo, S. Keita, M. Kinzer, F. K. George, B. Godefroy, G. Gutierrez, N. Kannangarage, H. L. Mills, T. Moller, S. Meijers, Y. Mohamed, O. Morgan, G. Nedjati-Gilani, E. Newton, P. Nouvellet, T. Nyenswah, W. Perea, D. Perkins, S. Riley, G. Rodier, M. Rondy, M. Sagrado, C. Savulescu, I. J. Schafer, D. Schumacher, T. Seyler, A. Shah, M. D. Van Kerkhove, C. S. Wesseh, Z. Yoti, Exposure Patterns Driving Ebola Transmission in West Africa: A Retrospective Observational Study. PLOS Med. 13, e1002170 (2016).

56. Stan Development Team, PyStan: the Python interface to Stan, Version 2.19.1.0. (2018).

57. B. W. Silverman, Density estimation for statistics and data analysis (CRC press, 1986), vol. 26.

58. P. Virtanen, R. Gommers, T. E. Oliphant, M. Haberland, T. Reddy, D. Cournapeau, E. Burovski, P. Peterson, W. Weckesser, J. Bright, S. J. van der Walt, M. Brett, J. Wilson, K. J. Millman, N. Mayorov, A. R. J. Nelson, E. Jones, R. Kern, E. Larson, C. J. Carey, I. Polat, Y. Feng, E. W. Moore, J. VanderPlas, D. Laxalde, J. Perktold, R. Cimrman, I. Henriksen, E. A. Quintero, C. R. Harris, A. M. Archibald, A. H. Ribeiro, F. Pedregosa, P. van Mulbregt, A. Vijaykumar, A. Pietro Bardelli, A. Rothberg, A. Hilboll, A. Kloeckner, A. Scopatz, A. Lee, A. Rokem, C. N. Woods, C. Fulton, C. Masson, C. Häggström, C. Fitzgerald, D. A. Nicholson, D. R. Hagen, D. V. Pasechnik, E. Olivetti, E. Martin, E. Wieser, F. Silva, F. Lenders, F. Wilhelm, G. Young, G. A. Price, G. L. Ingold, G. E. Allen, G. R. Lee, H. Audren, I. Probst, J. P. Dietrich, J. Silterra, J. T. Webber, J. Slavič, J. Nothman, J. Buchner, J. Kulick, J. L. Schönberger, J. V. de Miranda Cardoso, J. Reimer, J. Harrington, J. L. C. Rodríguez, J. Nunez-Iglesias, J. Kuczynski, K. Tritz, M. Thoma, M. Newville, M. Kümmerer, M. Bolingbroke, M. Tartre, M. Pak, N. J. Smith, N. Nowaczyk, N. Shebanov, O. Pavlyk, P. A. Brodtkorb, P. Lee, R. T. McGibbon, R. Feldbauer, S. Lewis, S. Tygier, S. Sievert, S. Vigna, S. Peterson, S. More, T. Pudlik, T. Oshima, T. J. Pingel, T. P. Robitaille, T. Spura, T. R. Jones, T. Cera, T. Leslie, T. Zito, T. Krauss, U. Upadhyay, Y. O. Halchenko, Y. Vázquez-Baeza, SciPy 1.0: fundamental algorithms for scientific computing in Python. Nat. Methods. 17, 261–272 (2020).

59. S. van Buuren, K. Groothuis-Oudshoorn, mice: Multivariate imputation by chained equations in R. J. Stat. Softw. 45, 1–67 (2011).

60. D. Bates, M. Mächler, B. M. Bolker, S. C. Walker, Fitting linear mixed-effects models using lme4. J. Stat. Softw. 67, 1–48 (2015).

61. W. N. Venables, B. D. Ripley, Modern Applied Statistics with S (Springer, New York, Fourth., 2002; http://www.stats.ox.ac.uk/pub/MASS4).

