## supplementary material for "Transmission heterogeneities, kinetics, and controllability of SARS-CoV-2"

**This PDF file includes:**

Materials and Methods
Figs. S1 to S11
Tables S1 to S5

### Materials and Methods

#### 1. Data source

#### 1.1 Epidemiological SARS-CoV-2 data

We collected data on 1,178 confirmed SARS-CoV-2 infections in Hunan Province, China, from January 16 to April 2, 2020, following a protocol for field epidemiological investigation developed by the National Health Commission of the People's Republic of China to identify potential COVID-19 cases (*52*). Primary and secondary SARS-CoV-2 infections were identified through: (i) active screening of incoming passengers into Hunan province and high-risk populations in the community who had a history of travel to Wuhan City/Hubei Province, capturing travel-associated symptomatic and asymptomatic infections; (ii) passive surveillance in hospitals and outpatient clinics, involving testing of individuals whose symptoms were compatible with COVID-19, capturing symptomatic cases; (iii) contact tracing of all confirmed infections identified by the above screening, followed by systematic monitoring of close contacts of these confirmed infections, capturing symptomatic and asymptomatic infections. All SARS-CoV-2 positive individuals in this database received positive laboratory confirmation of SARS-CoV-2 infection by RT-PCR test. Before February 7, 2020, contacts were tested if they developed symptoms during the quarantine period. After February 7, specimens were collected at least once from each contact during quarantine, regardless of symptoms. In total, 794 of the 1178 (67.8%) confirmed SARS-CoV-2 infections were diagnosed before February 7. Of the 794 infections detected before February 7$^{th}$, only 3% were asymptomatic. After February 7$^{th}$, the asymptomatic proportion among confirmed SARS-CoV-2 infections increased to 35.5%. A detailed flowchart of case ascertainment process is shown in Fig. S1.

The information collected for each case includes age, sex, prefecture (of case being reported), clinical severity (asymptomatic, mild, moderate, severe, or critical, see Table S1 for definition), potential exposures (travel history to Wuhan or contact with confirmed SARS-CoV-2 infection), time windows of potential exposures, date of the start of isolation/pre-symptomatic quarantine, date of symptom onset (list of symptoms below), date of healthcare consultation, date of hospital admission and ICU admission (if applicable), and date of laboratory-confirmation. The list of symptoms observed and documented among all patients includes: *fever (57.7%), dry cough (36.4%), fatigue (23.9%), sputum (19.6%), headache (10.3%), muscle ache (8.6%), sore throat (7.8%), chills (7.6%), chest tightness (5.4%), diarrhea (5.2%), shortness of breath (5%), runny nose (4.2%), stuffy nose (4.2%), vomiting (2.2%), joint pain (2.0 %), nausea (1.9%), breathing difficulty (1.4%), chest pain (1.3%), abdominal pain (0.5%), conjunctival hyperemia (0.3%)*. "Loss of taste/smell" was not included as a separate symptom as it was not known to be a symptom specific to COVID-19 at the time. However, if loss of taste/small had been reported by a patient it would have been included in the "other symptoms" category and used to estimate onset date along with other symptoms. All epidemiological information and testing data were collected by the Hunan CDC staff or by trained local CDC personnel and entered into a systematic database.

For each SARS-CoV-2 positive individual in the database, information was compiled on the start/end date of exposure, along with the dates of symptom onset (for symptomatic individuals) and laboratory confirmation. Biologically, the time of infection should occur before the onset of symptom or a positive RT-PCR test. Thus, we update the patient's end date of putative exposures in the database as the earliest of the reported exposure end date, date of symptom onset, or date of laboratory confirmation. If the start date of exposure is later than the date of symptom onset or positive RT-PCR test, it likely reflects recall error and we update the exposure start date as missing (1.9% of the records).

### 1.2 Contact tracing database

We collected data on 15,648 individuals in close contact with the 1,178 confirmed SARS-CoV-2 infections identified in Hunan Province based on the national protocol (*52*), representing 19,227 unique exposure events. Information included age, and sex of the contacts, type of contacts (household, extended family, social, community, and healthcare, see Table S2 for definition), as well as the start and end dates of contact exposure. If the contact was confirmed with SARS-CoV-2 by RT-PCR, a unique identifier mapping the individual to the SARS-CoV-2 patient database was provided.

Any individual reporting encounters as described in Table S2 and occurring within <1m of a SARS-CoV-2 infected individual (irrespective of displaying symptoms) was considered a close contact, at risk of SARS-CoV-2 infection. All records were extracted from the electronic database managed by Hunan Provincial Center for Disease Control and Prevention (*52*). All individual records were anonymized and de-identified before analysis.

### 1.3 Definition of a SARS-CoV-2 cluster

Based on the contact tracing database, we define a SARS-CoV-2 cluster as a group of two or more confirmed SARS-CoV-2 cases or asymptomatic infections with an epidemiological link, i.e. occurring through the same contact type (e.g. home, work, community, healthcare, or other) and for which a direct contact between successive cases can be established within two weeks of symptom onset of the most recent case (alternatively, the date of RT-PCR test for asymptomatic infections). In total, there are 210 clusters recorded in the database, for a total of 831 SARS-COV-2 infections.

While clusters of cases are grouped together based on shared exposures, a subset of cases report additional exposures outside the cluster as possible causes of infection as well. As a result, there can be more than one primary case within each cluster. In addition, for cases that only report exposures within the cluster, a unique infector cannot always be identified, given simultaneous SARS-CoV-2 exposures within the same cluster.

A sporadic case is defined as a laboratory-confirmed SARS-CoV-2 individual who does not belong to any of the reported clusters (i.e. a singleton who has no epidemiological link to other infections identified). In total, there are 347 sporadic cases recorded in the database.

Since the source and direction of transmission within a cluster cannot always be defined based on epidemiological grounds alone, we next turn to a modeling approach to probabilistically reconstruct infector-infectee transmission chains and further evaluate predictors of transmission.

### 2. Reconstruction of SARS-CoV-2 transmission chains

Reconstruction of transmission chains based on contact tracing data have been done with prior emerging outbreaks (*54*, *55*). In this section, we describe our sampling algorithm to stochastically reconstruct the transmission chains, which is customized to the unique contact tracing data of SARS-CoV-2 outbreaks in Hunan collected by the Hunan CDC and accounting for uncertainties in multiple plausible transmission routes compatible with the observation.

### 2.1 Sampling algorithm

For each cluster and each patient $i$ in the cluster, the time of infection $\mathrm{t}_{\mathrm{i}}^{\mathrm{inf}}$ is stochastically sampled by randomly drawing from the incubation period distribution and subtracting this value from the reported time of symptom onset, i.e. $\mathrm{t}_{\mathrm{i}}^{\mathrm{inf}} = \mathrm{t}_{\mathrm{i}}^{\mathrm{sym}} - \tau_{\mathrm{i}}^{\mathrm{incu}}$, where $\tau_{\mathrm{i}}^{\mathrm{incu}}$ is the sampled incubation period and $t_i^{sym}$ the date of symptom onset (*12*). The incubation period follows a Weibull distribution:

$$g_{incu}(\tau) = \frac{k}{\lambda}\left(\frac{\tau}{\lambda}\right)^{k-1} exp\left(-\left(\frac{\tau}{\lambda}\right)^{k}\right)$$

with shape parameter $k = 1.58$ and scale parameter $\lambda = 7.11$. The median incubation period is taken to be 5.56 days with IQR (3.14, 8.81) days (*12*).

The sampled time of infection $t_i^{inf}$ must satisfy the following constrains:

- $t_i^{inf}$ must fall within the start and end dates of the exposures identified by epidemiological investigation.
- For any infector-infectee pair, the time of infection of the infector $t_{infector}^{inf}$ must be earlier than the time of infection of the infectee $t_{infectee}^{inf}$, i.e. $t_{infector}^{inf} < t_{infectee}^{inf}$.

A SARS-CoV-2 infected individual may have multiple exposures (either through contacts with multiple SARS-CoV-2 infected individuals, or travel history to Wuhan in addition to contact with a SARS-CoV-2 individual). For an individual *i* who has multiple sources of exposure with a cluster, all other cases in contact with *i* are potential sources of infection, except for those whom *i* has infected. If the sampled infection time of infectee *i*, $t_i^{inf}$, satisfies the constraints of multiple exposures, we randomly choose one as the source of infection. If $t_i^{inf}$ satisfies the constrains of none of the plausible exposures, we resample $t_i^{inf}$ until individual *i* has one and only one valid source of infection. For individuals with missing onset dates (including all asymptomatic individuals), we set the time of infection as missing. The source of infection is then randomly chosen from all plausible exposures identified from epidemiological investigation.

We stochastically reconstruct 100 realizations of transmission chains to account for uncertainties in both the timing and source of exposures. 375 of the 831 (45%) SARS-CoV-2 infections do not have unique epidemiological link and their transmission routes may vary from one realization to another. In addition, only 35 of the 831 (4.2%) have missing onset dates.

We remove all singletons from the reconstruction of transmission chains, since they are not epidemiologically linked to other cases, but we consider these singletons when we analyze the distribution of secondary cases and when we represent the transmission network in Fig. 1.

#### 2.2 Distribution of the number of secondary infections among transmission chains

Next, we calculate the number of secondary infections for each of the 1,178 SARS-CoV-2 individuals based on the 100 reconstructed transmission chains among 831 cluster cases, and the 347 singletons. The distribution of secondary infections is shown in Fig. 1 We fit a negative binomial distribution to these data using package "pystan" version v2.19.1.1 (*56*) with uniform prior. We estimated mean $\mu = 0.40$, 95% CI 0.35 to 0.46 and dispersion parameter $k = 0.30$, 95%CI 0.23 to 0.39. In addition, we fit the geometric and Poisson distributions to the data. The negative binomial distribution best describes the data based on Akaike information criterion (Fig. 1).

### 3. Kinetics of SARS-CoV-2 transmission

#### 3.1 Generation interval and serial interval distribution

The generation interval is defined as the time interval between the dates of infections in the infector and the infectee. We calculate the generation intervals of all the infector-infectee pairs based on 100 realizations of the reconstructed transmission chains. The cumulative distribution of the generation interval is shown in Fig. S7A, C. The observed serial interval is defined as the time interval between dates of symptom onsets in the infector and the infectee. We calculate the serial interval of all the infector-infectee pairs based on 100 realizations of the reconstructed transmission chains with known dates of symptom onset. To further reduce potential recall bias on the timing of symptom onset/exposure, we down-sample the outlier incubation period pairs. To do this in a statistically sound manner, we rely on the independence of the incubation periods of the infector and the infectee, and down-sample infector-infectee pairs whose joint likelihood of the observed incubation period pair is very low. Specifically, we first estimate the joint empirical distribution of the incubation periods of both the infector and infectee using the gaussian kernel density estimate (*57*) in the package "scipy" version v1.5.0 function "scipy.stats.gaussian_kde" (*58*). The joint likelihood of observing the incubation periods of a given infector-infectee pair based on the kernel density estimate is denoted as $p_{kde}(\tau_i^{incu}, \tau_j^{incu})$. The joint likelihood of the incubation period of the same infector-infectee pairs based on two independent draws from the Weibull distribution $g_{incu}(\tau) = \frac{k}{\lambda}\left(\frac{\tau}{\lambda}\right)^{k-1} exp\left(-\left(\frac{\tau}{\lambda}\right)^k\right)$ with shape parameter $k = 1.58$ and scale parameter $\lambda = 7.11$ (Section 2.1) is denoted as $p_E(\tau_i^{incu}, \tau_j^{incu})$ . If $p_{kde}\left(\tau_i^{incu}, \tau_j^{incu}\right) > p_E\left(\tau_i^{incu}, \tau_j^{incu}\right)$, it suggests the observed incubation periods are over-represented relative to expectations, and vice versa. We introduce a down-sampling weight in accordance with the incubation period distribution as $w_{incu} = p_E\left(\tau_i^{incu}, \tau_j^{incu}\right)/p_{kde}\left(\tau_i^{incu}, \tau_j^{incu}\right)$. The distribution of the serial interval is shown in Fig. S7B, D.

**3.2 Gauging the impact of case isolation on the distribution of the serial and generation intervals.**

We select all infector-infectee pairs for which the infector has been isolated during the course of his/her infection, date of symptom onset is available, and times of infection have been estimated (range from 348 to 372 pairs across 100 sampled transmission chains). We stratify the data by the infector's time interval between onset and isolation, $\tau_{iso}$ , with $\tau_{iso} \in \{(-\infty, 2), [2, 4), [4, 6), [6, +\infty)\ days\}$, and assess how the generation interval and serial interval distributions change with the timeliness of case isolation (Fig. 3A and Fig. 3B). We use Mann-Whitney U test to compare the statistical significance in the differences of serial/generation interval distribution across different strata.

**3.3 Speed of case isolation and relative contribution of pre-symptomatic transmission.**

As cases are isolated earlier in the course of infection, we expect that the contribution of pre-symptomatic transmission will increase. This is because symptomatic transmission occurs after pre-symptomatic transmission and transmission will be blocked after effective isolation. In other words, isolated individuals remain infectious, but they can only effectively transmit before isolation, which is predominantly in their symptomatic phase. To validate the hypothesis that the contribution of pre-symptomatic transmission is affected by interventions, we first estimate the overall contribution of pre-symptomatic transmission among all reconstructed transmission chains. Let $tr_{i,j}^k$ represent each transmission event from an infector to infectee $i$, in realization $j$ of the 100 sampled transmission chains; $k = 0$ indicates that infection in an infectee occurred before the time of symptom onset of his/her infector, denoting pre-symptomatic transmission, while $k = 1$ indicates that the time of infection occurred after the infector's symptom onset (i.e. post-symptomatic transmission). Thus, the overall fraction of pre-symptomatic transmission in realization $j$ can be calculated using the following formula:

$$P_j^{pre} = \frac{\sum_i tr_{i,j}^{k=0}}{\sum_i \sum_k tr_{i,j}^{k}}$$

Mean and 95% CI of $P^{pre}$ can be estimated over the 100 realizations of the reconstructed transmission chains. We further stratify $P^{pre}$ by the time interval between an infector's symptom onset and isolation, considering four categories (days):

$$(-\infty,0),\ [0,2),\ [2,4),\ [4,6),\ [6,+\infty)$$

The mean and variance (based on 100 realizations of the sampled transmission chains) of $P^{pre}$ for each category of the isolation intervals is shown in Fig. 3C.

**3.4 Relative infectiousness profiles over time adjusted for case isolation.**

In Hunan province, all COVID-19 cases regardless of clinical severity were managed under medical isolation in appointed hospitals; while contacts of SARS-CoV-2 infections were quarantined in designated medical observation centers. In Section 4, we estimate that the risk of transmission through healthcare contacts is the lowest among all contact types, thus case isolation and contact quarantine are highly effective to block onward transmission after isolation/quarantine. As a result, the observed serial/generation intervals are shorter than they would be in the absence of case isolation and contact quarantine. The censoring effects are clearly demonstrated in Fig. 3A and Fig. 3B, where we observe that the median generation time drops from 7.1 days for $\tau_{iso} > 6\ (days)$ after symptom onset, to 4.0 days for $\tau_{iso} < 2\ (days)$.

Moreover, the timeliness of case isolation is not static over time. Fig. S8 shows the distribution of time from symptom onset to isolation in three different phases of epidemic control (*Phase I, II, and III*) defined by two major changes in COVID-19 case definition issued by National Health Commission on Jan. 27 and Feb. 4. The median time from symptom onset to isolation decreases from 5.4 days in *Phase I* to -0.1 days in *Phase III*, due to the expansion of "suspected" case definition (*51*) and strengthening of contact tracing effort (Fig. S8).

**3.4.1 Generation interval adjusted for case isolation.**

Estimating the generation interval distribution in the absence of interventions is important to understand the kinetics of SARS-CoV-2 transmission, as the shape of the generation interval distribution represents the population-average infectiousness profile since the time of infection. To minimize the potential error of flipping the directionality of infector-infectee relationship during contact tracing, we further limit our analysis to the infector-infectee pairs where the primary case had a travel history to Wuhan (and no other SARS-CoV-2 contact), while the secondary case did not have a travel history to Wuhan but was epidemiological linked to the primary case. To further reduce potential recall bias on the timing of symptom onset/exposure, we down-sample the outlier incubation period pairs. To do this in a statistically sound manner, we rely on the independence of the incubation periods of the infector and the infectee, and down-sample infector-infectee pairs whose joint likelihood of the observed incubation period pair is very low. Specifically, we first estimate the joint empirical distribution of the incubation periods of both the infector and infectee using the gaussian kernel density estimate (*57*) in the package "scipy" version v1.5.0 function "scipy.stats.gaussian_kde" (*58*). The joint likelihood of observing the incubation periods of a given infector-infectee pair based on the kernel density estimate is denoted as $p_{kde}(\tau_i^{incu}, \tau_j^{incu})$. The joint likelihood of the incubation period of the same infector-infectee pairs based on two independent draws from the Weibull distribution $g_{incu}(\tau) = \frac{k}{\lambda}\left(\frac{\tau}{\lambda}\right)^{k-1} exp\left(-\left(\frac{\tau}{\lambda}\right)^{k}\right)$ with shape parameter $k = 1.58$ and scale parameter $\lambda = 7.11$ (Section 2.1) is denoted as $p_E(\tau_i^{incu}, \tau_j^{incu})$. If $p_{kde}(\tau_i^{incu}, \tau_j^{incu}) > p_E(\tau_i^{incu}, \tau_j^{incu})$, it suggests the observed incubation periods are over-represented relative to expectations, and vice versa. We introduce a resampling weight in accordance with the incubation period distribution as $w_{incu} = p_E(\tau_i^{incu}, \tau_j^{incu})/p_{kde}(\tau_i^{incu}, \tau_j^{incu})$. The resampling weights as a function of the incubation periods among the infector and infectee are visualized in Fig. S11.

To account for the "censoring" of generation interval distribution due to quarantine/case isolation, we first exclude generation intervals where transmission occurred after isolation of the infector (only 4.3% of the reconstructed transmission events, attesting to the effectiveness of isolation). We then divide the generation intervals into three groups based whether the date of symptom onset of the infectors fall within a given phase of epidemic control in Hunan. In Group 1 the illness onset of the infectors occurred before Jan. 27th (*Phase I*); in Group 2 the illness onset of the infector occurred between Jan. 27th and Feb. 4th (*Phase II*); in Group 3, the illness onset of the infector occurred after Feb. 4th (*Phase III*). For a given generation interval $\tau_{GI}$ of an infector-infectee pair in each group, we denote:

- The time of symptom onset of the infector as $t_{onset}$.
- The time of case isolation/quarantine of the infector as $t_{iso}$.
- The time of transmission from the infector to the infectee as $t_{inf.}$.
- The time interval between onset of the infector and transmission to the infectee $\tau_{oi} = t_{onset} - t_{inf.}$
- The time interval between infection times in the infector and infectee, i.e. the generation interval $\tau_{ii}$
- The probability distribution from symptom onset to isolation as $P_i(\tau_{iso})$, where $i \in \{I, II, III\}$ denotes the different phases of epidemic control, determined by symptom onset in the infector $t_{onset}$. The functional form of $P_i(\tau_{iso})$ is shown in Fig. S8. The corresponding cumulative probability distribution is denoted as $C_{OI}^{i}(\tau_{iso})$.

The probability of this infection-infectee pair escaping the "censoring" due to quarantine and case isolation is $p_i^{esc.} = 1 - C_{OI}^{i}(\tau_{oi})$. For every n observations of the generation interval $\tau_{ii}$ under intervention $p_i(\tau_{iso})$ given $\tau_{oi}$, there should be $m = \frac{n}{p_i^{esc.}}$ observations of $\tau_{GI}$ given $\tau_{oi}$ without intervention $p_i(\tau_{iso})$. Thus, we denote the sampling weight adjusted for case isolation as $w_{iso} = \frac{1}{p_i^{esc.}}$. The overall resampling weight of generation interval $\tau_{ii}$ between infector $i$ and infectee $j$ considering both incubation period distribution and censoring due to case isolation is given by $w_{sample}(i,j) = w_{incu} \times w_{iso} = \frac{p_E(\tau_i^{incu}, \tau_j^{incu})}{p_i^{esc.} \times p_{kde}(\tau_i^{incu}, \tau_j^{incu})}$. We resample from $\{\tau_{ii}(i,j)\}$ with sampling weights $w_{sample}(i,j)$ until we reach a sample size of $n = 100$ to obtain the distribution of generation time $\{\tau_{ii}^{adj.}\}$ adjusted for censoring. The distribution of $\tau_{ii}^{adj.}$ reflects the generation interval that would have been observed in the absence of quarantine and case isolation/quarantine. We fit Weibull, gamma, and lognormal function to $\{\tau_{ii}^{adj.}\}$. The distribution of $\tau_{ii}^{adj.}$ is best described by the Weibull distribution:

$$g_{GI}^{adj.}(\tau) = \frac{k}{\lambda}\left(\frac{\tau}{\lambda}\right)^{k-1} exp\left(-\left(\frac{\tau}{\lambda}\right)^{k}\right)$$

with $k = 1.60$ and $\lambda = 6.84$ (Fig. S10).

**3.4.2 Distribution of time interval between symptom onset and transmission, adjusted for case isolation.**

In contrast to the generation interval distribution, which characterizes the relative infectiousness of a SARS-CoV-2 infection over time with respect to the time of infection, we now focus on the interval between symptom onset and transmission. This shifts the reference point of the infectiousness profile from the time of infection to the time of symptom onset. Namely the distribution of symptom onset to transmission adjusted for case isolation $\{\tau_{OT}^{adj.}\}$ represents the population-average relative infectiousness profile over time since the onset of symptom. Of note, since we observe substantial pre-symptomatic transmission for SARS-CoV-2, negative values of $\tau_{OT}^{adj.}$ are allowed.

Similarly to the previous section, we resample from $\{\tau_{OT}(i,j)\}$ with sampling weights $w_{sample}(i,j)$ until a sample of size $n = 100$ is reached to obtain the distribution of symptom onset to transmission $\{\tau_{OT}^{adj.}\}$. The resampled distribution represents the infector's relative infectiousness (population average) with respect to the infector's symptom onset (Fig. S6B). The best-fit distribution is a normal distribution:

$$f_{OT}^{adj.}(\tau_{OT}^{adj.}) = \frac{1}{\sigma\sqrt{2\pi}} e^{-\frac{1}{2}\left(\frac{\tau_{OT}^{adj.}-\mu}{\sigma}\right)^2}$$

with mean $\mu = -0.24$ days and standard deviation $\sigma = 3.40$ days. After adjusting for case isolation, the fraction of transmission occurring during the pre-symptomatic phase of SARS-CoV-2 infection is 54%, (95%CI 38%, 0.70%).

**3.5 Estimating the basic reproduction number in Wuhan before lockdown**

A recent study (*33*) estimated the initial growth rate of the epidemic in Wuhan at 0.15 day$^{-1}$ 95% CI (95% CI, 0.14 to 0.17) ahead of the lockdown. The estimate is based on the daily rise in reported cases by onset date; adjustment for increased reporting due to a broadening case definition places the growth rate at 0.08 day$^{-1}$ (*33*). The Euler–Lotka equation (*34*) describes the relationship between the basic reproduction number $R_0$, the epidemic growth rate $r$, and the generation interval distribution $g(\tau)$:

$$R_0 = \frac{1}{\int exp(-r * \tau) \times g(\tau) d\tau}$$

We assume that no effective intervention had been implemented in Wuhan by the time of the lockdown (Jan. 23). Using the generation time distribution adjusted for "censoring" due to quarantine and case isolation $g_{GI}^{adj.}(\tau)$ described in the previous section, we estimate the basic reproduction number in Wuhan during the exponential growth phase at $R_0^{Wuhan} = 2.17$, (95% CI, 2.08 to 2.36), based on the conservatively higher estimate of growth rate in this city (0.15 day$^{-1}$ 95% CI (95% CI, 0.14 to 0.17). If we rescale the adjusted generation time distribution $g_{GI}^{adj.}(\tau)$ by a factor of $R_0^{Wuhan}$, the function

$$r_{GI}(\tau) = g_{GI}^{adj.}(\tau) \times R_0^{Wuhan}$$

represents the average risk of SARS-CoV-2 transmission to a secondary case at time $\tau$ since infection. The red line in Fig. 3D visualizes the functional form of $r_{GI}(\tau)$.

Similarly, if we rescale the adjusted distribution of symptom onset to transmission $f_{OT}^{adj.}(\tau)$ with $R_0^{Wuhan}$, the function

$$r_{OT}(\tau) = f_{OT}^{adj.}(\tau) \times R_0^{Wuhan}$$

represents the average risk of transmission to a secondary case at time $\tau$ since the symptom onset of the infector. The red line in Fig. 3E visualizes the functional form of $r_{OT}(\tau)$.

**3.6 Evaluating the impact of case isolation and quarantine on SARS-CoV-2 transmission.**

To evaluate the impact of quarantine and case isolation on the reduction of SARS-CoV-2 transmission at different phases of epidemic control, we denote the time intervals between a patient's time of infection to his/her time of isolation as $\tau_{inf}^{iso}$. The corresponding probability distribution is $p_{ii}^{j}(\tau)$, where $j \in \{I, II, III\}$ denotes the phase of epidemic control. We denote the distribution of the incubation period $\tau_{incu}$ as $p_{incu}(\tau)$ and the distribution of symptom onset to isolation $\tau_{onset}^{iso}$ as $p_{oi}^{j}(\tau)$, for each phase $j$ of epidemic control. We sample $\tau_{inf}^{iso} = \tau_{incu} + \tau_{onset}^{iso}$ numerically through independently sampling of $\tau_{incu}$ and $\tau_{onset}^{iso}$ and add them together. Fig. S9 shows the distribution of 100 numerical sampling of $\tau_{inf}^{iso}$ at different phases of epidemic control. We fit the sampled distribution of $\tau_{inf}^{iso}$ to various probability distributions including normal, lognormal, gamma, Cauchy, logistic, and hyperbolic secant distribution. The top three fits are show in Fig. S9 and the best fit is selected based on the Akaike information criterion during each of the three phases of epidemic control. We denote cumulative density distribution of $p_{ii}^{j}(\tau)$ as

$$C_{ii}^{j}(\tau) = \int_{-\infty}^{\tau} P_{ii}^{j}(\tau')d\tau'$$

where $C_{ii}^{j}(\tau)$ gives the probability that transmission is blocked after time $\tau$, where $\tau$ is the time since infection. The shaded areas in Fig. S9 visualize the probabilities $C_{ii}^{j}(\tau)$ for the best-fit distribution.

Assuming that all SARS-CoV-2 patients are subject to case isolation and quarantine efforts carried out in Hunan province, we can estimate the average risk of transmission $r_{GI}^{control(j)}(\tau)$ of an infected individual at time $\tau$ since his/her infection, during phase $j \in \{I,\ II,\ III\}$ of epidemic control as:

$$r_{GI}^{control(j)}(\tau) = r(\tau) \times \left(1 - C_{ii}^{j}(\tau)\right) = g_{GI}^{adj.}(\tau) \times R_0^{Wuhan} \times \left(1 - C_{ii}^{j}(\tau)\right)$$

The corresponding basic reproduction number assuming 100% SARS-CoV-2 infection detection rate is given by:

$$R_0^{j} = \int_{0}^{\infty} r_{GI}^{control(j)}(\tau)\, d\tau\, , j \in \{I, II, III\}$$

In Fig. 3D, we visualize the transmission profile with respect to infection time $r_{GI}^{control(j)}$ for all three phases of epidemic control (dashed lines) and shows the estimated values of the corresponding basic reproduction number $R_0^{j}$.

Similarly, following Section 3.4.1, $C_{oi}^{j}(\tau)$ gives the probability that transmission is blocked after time $\tau$ since symptom onset in the infector, for the 3 phases of epidemic control $j \in \{I,\ II,\ III\}$. We can estimate the average risk of transmission $r_{OT}^{control(j)}(\tau)$ of an infected individual at time $\tau$ since his/her onset of symptom, during phase $j \in \{I,\ II,\ III\}$ of epidemic control as:

$$r_{OT}^{control(j)}(\tau) = r(\tau) \times \left(1 - C_{oi}^{j}(\tau)\right) = f_{OT}^{adj.}(\tau) \times R_0^{Wuhan} \times \left(1 - C_{oi}^{j}(\tau)\right)$$

In Fig. 3E, we visualize the transmission profile with respect to symptom onset time $r_{OT}^{control(j)}$ for all three phases of epidemic control (dashed lines).

#### 3.7 Evaluating synergistic effects of individual-level and population-level interventions on SARS-CoV-2 transmission.

We start by characterizing the controllability of SARS-CoV-2 (measured as $R_0$ under control measures) as a function of infection isolation rate and the speed of case isolation/pre-symptomatic quarantine. In Fig. 3F, we plot the phase diagram of $R_0$ as a function of infection detection proportion (fraction of all SARS-CoV-2 infections detected) and the mean time from symptom onset to isolation/quarantine $\tau_{iso}$. Contour lines indicates reductions in R0 from baseline non-intervention conditions. It is worth noting that we do not know the precise prevalence of truly asymptomatic infections as well as their role in transmission. Here we assume that asymptomatic cases have a similar shape of infectiousness profile over the course of infection as symptomatic cases, and a peak of infectiousness corresponding to the time of symptom onset in symptomatic cases, as shown in Fig. S10. The corresponding $\tau_{iso}$ for asymptomatic cases is measured as time from peak infectiousness to isolation. Here we assume that the distribution of symptom onset/peak infectiousness to isolation follows a normal distribution with mean $\tau_{iso}$ and standard deviation of 2 days.

We further consider the synergic effects of layering individual-based intervention (case isolation, contact tracing, and quarantine) with population-based interventions (i.e., via physical distancing, measured as a reduction in effective contact rates). In Fig. 3G, we plot the phase diagram of $R_0^E$ as a function of the proportion of population-level contact reduction and infection isolation rate, with the average speed of isolation 0 days after symptom onset/peak infectiousness and standard deviation of 2 days. The base $R_0 is$ 2.19, which is compatible with a growth rate of 0.15 observed in Wuhan without the adjustment for change in reporting (Section 3.5). The blue area indicates the region below the epidemic threshold, where control is achieved, and the red area indicates the region above the epidemic threshold. The phase diagram shows the effect of ramping up population-based interventions (i.e. increasing percent reduction in the effective contact rate) and strengthening individual-based interventions (i.e. increasing fraction of active infections identified and isolated). Both types of interventions act synergistically to reduce the effective reproduction number and consequently slow down the transmission. The dashed line in Fig. 3G indicates the minimum level of individual and population-based interventions required to stop transmission (i.e. bring $R_0^E = 1$), separating the regime of controlled and uncontrolled epidemics. It also demonstrates the trade-off between individual and population-based interventions: expanding efforts in case detection and isolation will reduce the amount of population-level interventions required to maintain control of the epidemic and vice versa.

Last, we consider a sensitivity analysis of lower baseline transmission scenario with base $R_0 = 1.57$, using a growth rate of $r = 0.08$, as observed in Wuhan data after adjustment for changes in reporting (Section 3.5). In Fig. 3H, we plot the phase diagram of $R_0$ as a function of proportion of population-level contact reduction (i.e. through physical distancing) and isolation rate, assuming that SARS-CoV-2 infections are isolated 2 days after symptom onset/peak infectiousness on average with a standard deviation of 2 days. Overall, the phase diagram bears similar interpretations as Fig. 3G which represents the scenario where $R_0 = 2.19$ and infections are isolated immediately upon symptom onset or at peak infectiousness. With a lower base reproduction number of 1.57, the requirements for both individual and population-based interventions to achieve control are lower. For instance, with a "relaxed" timeliness of isolation 2 days after symptom onset on average, control can be achieved by detection and isolation of 40% of all infections, associated with a reduction of 25% of effective contacts through population-level interventions. In contrast, in the high baseline transmission scenario (Fig. 3G), a 25% reduction in effective contacts needs to be coupled with an 80% infection detection and isolation, promptly upon symptom presentation, to achieve control. This is a much higher bar when compared to the low baseline transmission scenario.

### 4. Evaluating individual-level transmission heterogeneity of SARS-CoV-2

#### 4.1 Regression analysis to evaluate the "per-exposure" risk of SARS-CoV-2 transmission as a function of demographical, epidemiological, clinical, and behavioral predictors.

In this section, we use a mixed effects multiple logistic regression model to evaluate the risk of SARS-CoV-2 transmission for each exposure reported in the contact tracing database. we analyze the infection risk among a subset of 14,622 individuals who were close contacts of 870 SARS-CoV-2 patients. This dataset excludes primary cases whose infected contacts report a travel history to Wuhan, to avoid confounding in the source of infection. The dataset represents 74% of all SARS-CoV-2 cases recorded in the Hunan patient database. Each entry in the database represents a contact exposure between a SARS-CoV-2 infected individual and his/her contact. For individuals who were in contact with SARS-CoV-2 infected individual, the contact individual's age, sex, type of contact, the start/end dates of exposure, as well as the infection status (whether the exposed individuals was eventually infected with SARS-CoV-2) are carefully documented (Section 1.2). All SARS-CoV-2 infected individual (both primary cases and secondary infections via contact exposures) have unique identifiers that can be mapped to the SARS-CoV-2 patient line-list database, where additional information about the course of infection is also available (see Section 1.1 for detailed information). An individual in the contact tracing database can be exposed to multiple SARS-CoV-2 cases; further, an individual in the contact tracing database can be exposed to the same SARS-CoV-2 case through multiple independent exposures. All exposures are recorded independently.

For each exposure in the contact-tracing database, the regression outcome is coded as 1 if the contact eventually becomes infected and 0 if not infected. For each exposure, a list of independent variables, their definitions, and corresponding values are shown in Table S3 (fixed effects in the mixed model).

We also introduce random effects for each SARS-CoV-2 case, representing the individual-level infectiousness heterogeneity that is not explained by the independent variables representing fixed effects. These random effects also take into account the lack of independence of our observations.

A contact could report more than one SARS-CoV-2 exposure. If the contact eventually becomes infected, however, only one of the many exposures will be the actual source of infection. In this case, if we denote the number of exposures as $n^{expo}$, for each of the contact's $n^{expo}$ exposure entries in the database with two different outcomes, either the contact became infected (1 as regression outcome) with regression weight $1/n^{expo}$ or the contact avoided infection from the same exposure (0 as regression outcome) with regression weight $(n^{expo} - 1)/n^{expo}$. We exclude primary cases whose infected contacts report a travel history to Wuhan, as the infection could possibility originate from exposures in Wuhan in addition to exposure to local cases in Hunan.

A fraction of the regression variables has missing values in the contact-tracing database (see Table S3, column 3). We adopted the state-of-the-art "Multivariate Imputation by Chained Equations" algorithm (*59*) (implemented in R package "MICE" version 3.9.1 https://cran.r-project.org/web/packages/mice/index.html) to impute missing values in the database. All independent variables in Table S3 are used as predictors for data imputation. The number of multiple imputations is set as 10 with each imputation running 10 realizations. For each of the 5 realizations of imputed contact-tracing databases, we independently perform mixed effects multiple logistic regression of the risk of SARS-CoV-2 transmission with all exposures and variables described in Table S3 as covariates. The regression is performed using R package "lme4" (*60*) version v1.1-23 function "glmer" (https://cran.r-project.org/web/packages/lme4/index.html). The final odds-ratio estimates are pooled from the 5 independent regressions on 5 imputed databases using "MICE" package's "pool" function, based on Rubin's rule (*59*). The point (maximum likelihood) estimates of the odds ratios of independent variables, their 95%CIs, and the baseline odds (intercept) are reported in Fig. S3A.

To examine the model's fit to the data, we explore (i) how well the model reproduces the age profiles of infector-infectee pairs and (ii) whether the model captures the amount of transmission that occurs through different contact types (household, family, transportation, etc). We first randomly choose one of the five imputed contact-tracing databases. For each exposure entry in the imputed contact-tracing database, we calculate the model predicted risk of infection based on all fixed variables in the regression. We simulate the infection status of the contact according to the predicted risk by drawing from a binomial distribution. We repeat the process for all contacts, and further simulate 100 realizations of projected infection databases to gauge variability. Fig. S3C shows the observed age distribution of the infector-infectee pairs in the original data, and Fig. S3B visualizes the projected age distribution based on the regression model, averaged over 100 realizations. Violin plots in Fig. S3D show the relative fraction (with projection uncertainties) of transmission that is explained by each type of contacts, based on the model, while the dots in Fig. S3D represents the empirical observations. We find that the model accurately captures the strong assertiveness of transmission in the 30-50 years age group, and the off diagonals that represent transmission between different generations. Further, the model reproduces the relative contribution of different types of contacts seen in the empirical data (Fig. S3D).

### 4.2 Sensitivity analyses

#### 4.2.1 On contact type categories

We use sensitivity analyses to answer two questions related to the role of contacts (i) Are the 5 categories of contacts, as defined in the main analysis and broken down by timing of exposure, the most parsimonious way to explain transmission risk? This is especially important as there is overlap in the 95% CIs of the odds ratios associated with each type of contact. We can test this by collapsing some of the contact categories, and/or collapsing contacts of the same type over timing of exposure and assessing model fit. (ii) Does contact duration have similar effect on different types of contacts? In the reference model, we used duration as an independent variable that modulates the baseline risk of each contact type. But what if duration has a differential effect on each contact type? To address this question, we can incorporate the additional interaction between contact type and duration to the regression and assess model fit.

To test hypothesis (i), we fit the data to 5 other candidate regression models M1, M2, M3, M4, and M5, collapsing contact categories and time. We compare models M1-M5 to the reference model (M0) of the main analysis, while keeping other predictors of the regression unchanged. We find that M0 has the lowest AIC scores of all candidate models, indicating that distinguishing the timing of exposure as well as contact type best explains the data (Table S4).

Another sensitivity analysis (M6) was performed to examine whether including the interaction term between contact types and duration of contacts improves model fit. Our baseline model (M0) still best explains the data based on Akaike information criterion (Table S4).

##### 4.2.2 On missing data and change in testing protocols

As a sensitivity analysis for missing data imputation (especially addressing the issue of imputing "onset within exposure" for SARS-CoV-2 infected individuals that are asymptomatic), we perform a GLMM-logit regression with entries of missing data removed. To explore transmissibility from younger and older children, we further break-up the age bracket of predictor "Age (case)" (Table S3) into ***0-12 years***, ***12-25 years***, ***26-64 years***, and ***65+ years***. We remove the predictor of "onset within exposure", however, for predictor "clinical severity (case)", we break down the category "mild & moderate", and "severe & critical" based on whether the onset of the primary case occurred within the exposure time window. "(-)" indicate symptom onset outside the exposure time window, while "(+)" indicate symptom onset within the exposure time window. There was a change in the testing protocol for close contacts during the outbreak: prior to February 7, only contacts displaying symptoms were tested, while after February 7 all contacts were tested regardless of symptoms (Section 1.1). To evaluate the impact of the change in testing protocol before and after February 7 on age-dependent susceptibility, we further stratify the age groups of the contacts by the date of diagnostic of the corresponding primary (before/after February): 4 age groups (***0-12***, ***12-25***, ***26-64***, ***65+ years***) whose primary cases were diagnosed prior to February 7 and four additional age groups (***0-12***, ***12-25***, ***26-64***, ***65+ years***) whose primary cases were diagnosed after February 07, resulting a total of 8 age groups for the contacts. The results of the regression are shown in Fig. S4. Compared, to the main analysis in Fig S3, we see that the gradient in susceptibility is preserved in both time periods. Further, we find odds ratio (OR) for susceptibility among children under 12 years (relative to 26-64 years) in the data after February $7^{th}$ is comparable with the main analysis (OR=0.3 with 95% CI 0.11 to 0.86 v.s. OR=0.41 with 95% CI 0.26 to 0.63 in the main analysis).

#### 4.3 Regression analysis evaluating predictors of individual contact patterns among SARS-CoV-2 cases and the impact of interventions

While Section 4.1 addresses predictors of "per-contact" transmission risk heterogeneity, in this section we aim to characterize variation in individual contact patterns of SARS-CoV-2 cases by type of contact. We are particularly interested in the impact of both individual-based and population-based intervention on contact rates. Intuitively, the overall transmission rate of an infectious individual can be interpreted as the sum of contact rates across contact categories weighted by the "per-contact" transmission risk. Thus, conditioning on all other predictors, higher contact rates would translate to higher transmission rates.

We use regression analysis to model the individual contact patterns of each symptomatic SARS-CoV-2 case, whose contacts are traced and documented in the contact-tracing database. We focus on symptomatic cases (the majority of our data) because we are particularly interested in contacts near the time of symptom onset, since we have previously shown that transmission risk is highest near symptom onset. We first define a time window $\tau_{symp.}$ of peak infectiousness as $\pm 5$ days before and after each case's symptom onset $t_{symp.}$. This time window accounts for a majority (87%) of the total infection risk of a typical symptomatic SARS-CoV-2 infection (Fig. S10B, D). In addition, we consider the 4 main contact types separately: community, social, family, and household contacts. For each symptomatic SARS-CoV-2 case and contact type $s$, we denote the number of contacts on day $i$ as $k_i^s$. Here each contact in $k_i^s$ is weighted by the regression odds ratios of GLMM-logit, excluding effects from duration of exposure and if onset is within exposure time window. The cumulative daily contact rate $CCR_{\tau_{symp.}}^s$ within the time window $\tau_{symp.}$ for a given case is given by:

$$CCR_{\tau_{symp.}}^s = \sum_{i=t_{symp.}-5}^{t_{symp.}+5} k_i^s \times w(t_{symp.} - i)$$

Here $w(\tau)$ is the infectiousness profile with respect to symptom onset (Fig. S10B, D). Clearly, case isolation will impact an infected individual's contact rate, irrespective of whether the case is symptomatic. However here we restrict our analysis to symptomatic cases as the speed of case isolation and pre-symptomatic quarantine can be quantitatively measured as the time from isolation/pre-symptomatic quarantine to symptom onset.

To quantify the impact of socio-demographic factors and interventions on $CCR_{\tau_{symp.}}^s$, we consider a negative binomial regression with $CCR_{\tau_{symp.}}^s$ as the dependent variable and proxies of interventions intensities as independent variables in the regression. Specifically, we use a within-city mobility index as a proxy for the intensity of population-level social distancing, while we use time between isolation and symptom onset to measure the intensity of individual-level interventions (here, case isolation). We also include demographic and clinical predictors as independent variables to adjust for age and sex differences, as well as other changes in contact patterns. A full description of all regression variables is shown in Table S5.

The regression is performed using the R package "MASS" (*61*) version 7.3-51.6 function "glm.nb" (https://cran.r-project.org/web/packages/MASS/index.html). The point (maximum likelihood) estimates rate ratios along with their 95% CIs for each of the variables are presented in the bottom panels of Fig. 2E. We identify an effect of interventions on contact rates, along with clinical factors; these effects tend to be most intense for social and community contacts.

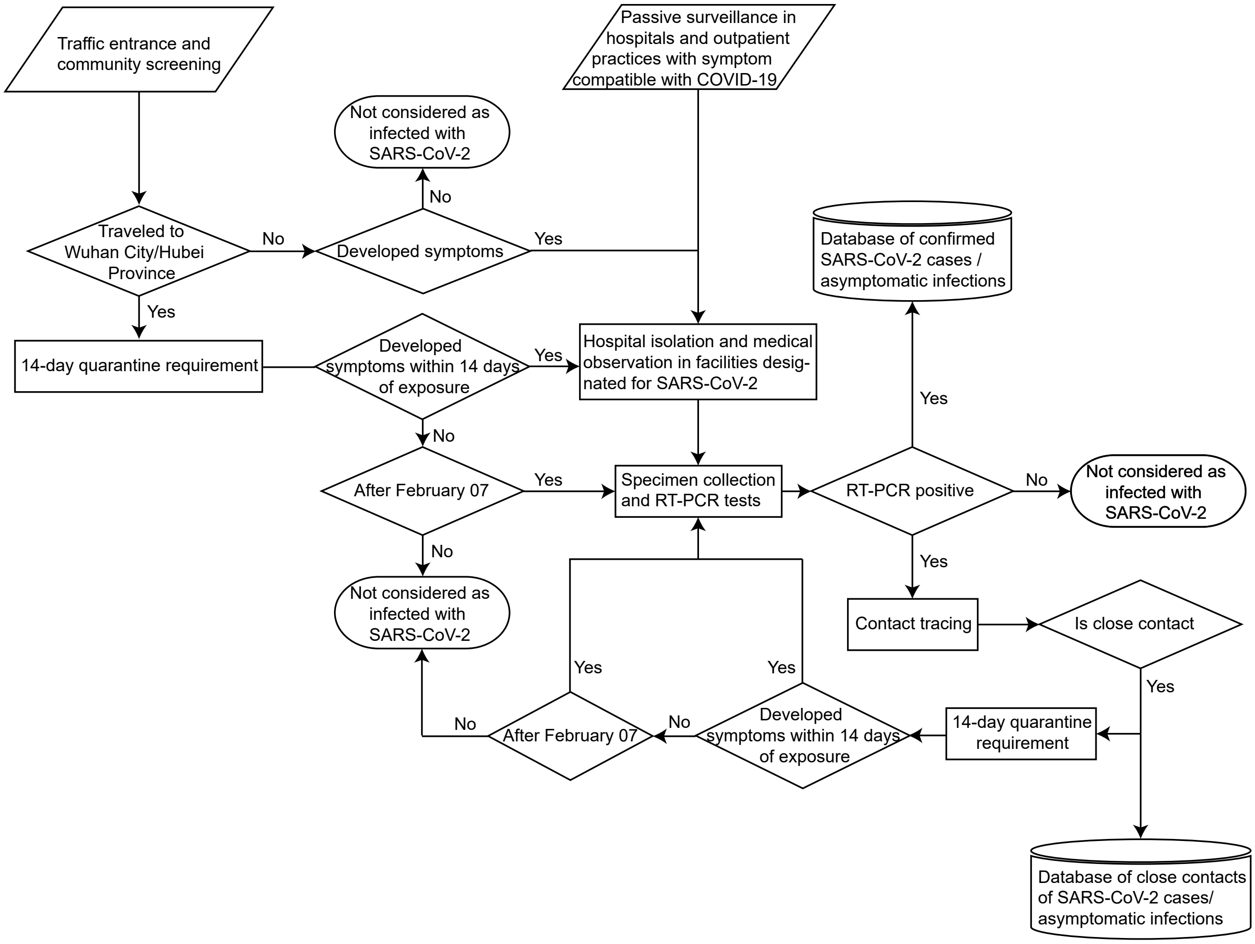


**Fig. S1.** Flowchart of SARS-CoV-2 cases/asymptomatic infection ascertainment process.

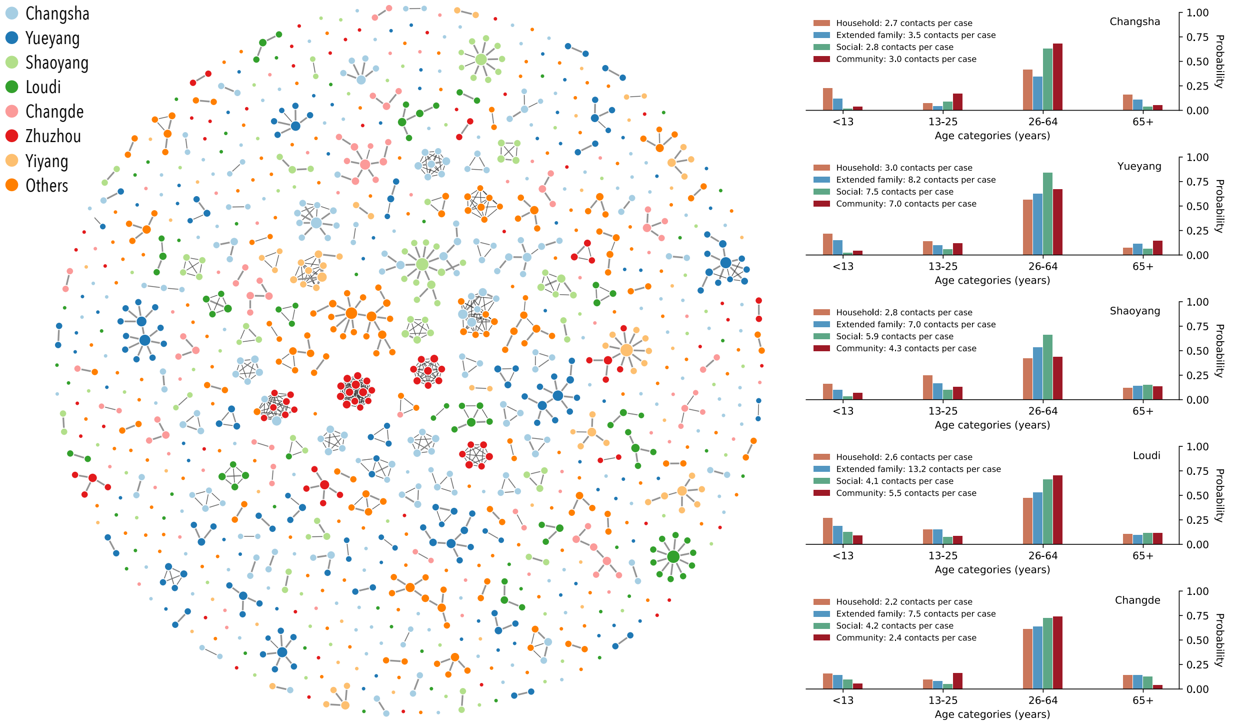


**Fig. S2. The topological uncertainties of transmission chains reconstruction and spatial variation of contact patterns.** Left: The network of the aggregation of 100 sampled transmission chains. Each node in the network represents a patient infected with SARS-CoV-2 and each link represents an infector-infectee relationship. The weights (visualized as widths) of the links are proportional to the probability of occurrence among 100 samples. Colors of the node denote the reporting prefecture of infected individuals. Right: The variation of contact patterns of the top 5 prefectures in Hunan with most SARS-CoV-2 infections, based on the contact tracing database. Bar plots are the age distribution of close contacts of SARS-CoV-2 infected individuals, across 4 age groups (<13 years, 13-25 years, 26-64 years, and >65 years), and stratified by contact types, based on the contact tracing database. Legends also reported the average number of close contacts of a SARS-CoV-2 infection in each of the 4 contact types.

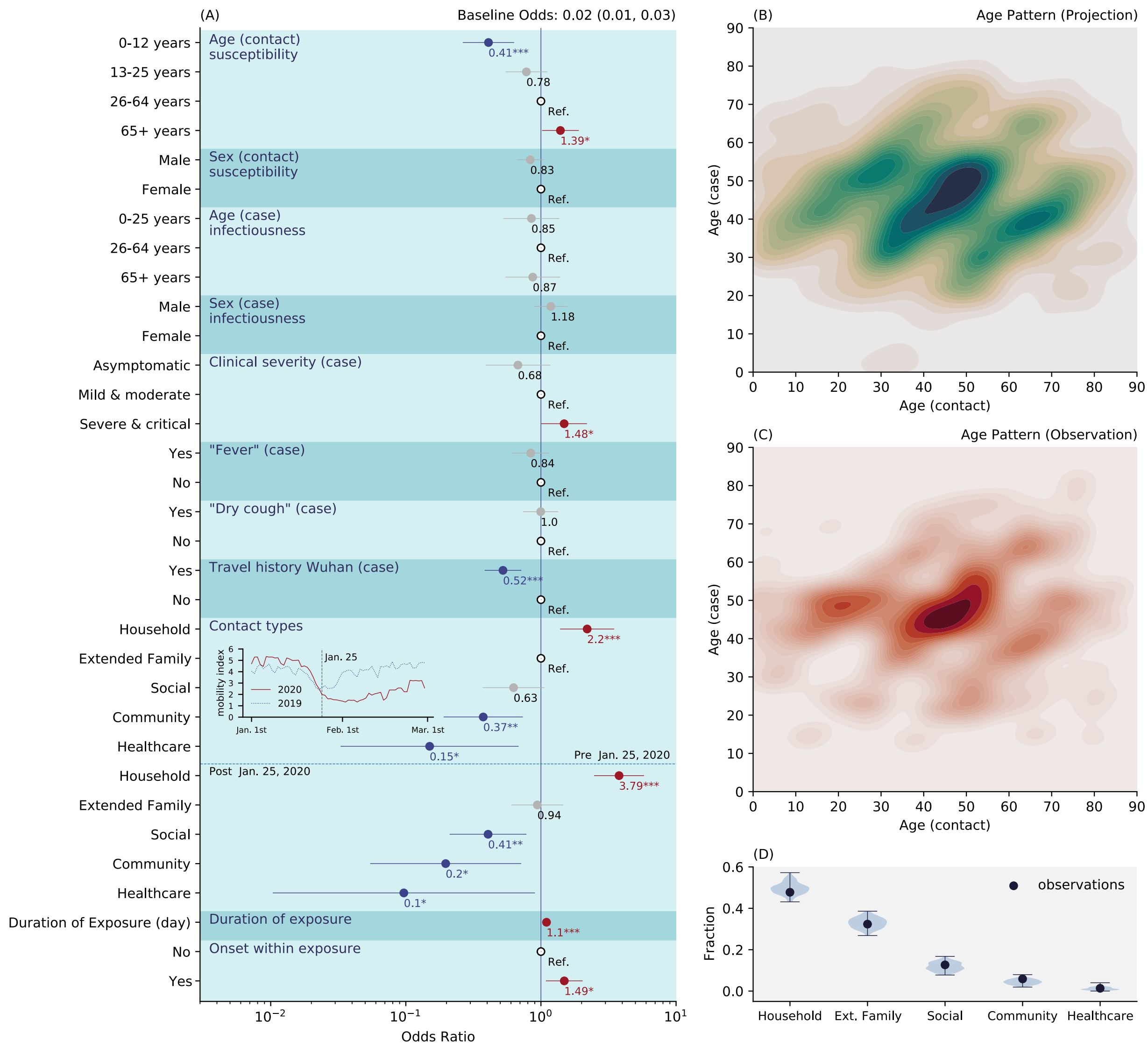


**Fig. S3.** (A) Individual predictors of transmission risk among close contacts of SARS-CoV-2 infected indivdiuals in Hunan. The predictors of the logistic regression as those indicated on the left (fixed effects) and we also include random effects for individual SARS-CoV-2 infections. Dots and lines indicate point estimates and 95% confidence interval of the odds ratio, numbers below the dots indicate the numerical value of the point estimates; "Ref." stands for reference category; * indicates p-value<0.05, ** indicates p-value<0.01, *** indicates p-value<0.001. Top inset indicates the within city mobility index in Changsha, Hunan for year 2020 and 2019, provide by Baidu Qianxi (*25*); dashed line indicates January 25, 2020. (B) Age distribution of projected infector-infectee pairs based on the regression model (average over 100 ensemble projections). (C) Age distribution of observed infector-infectee pairs. (D) The contribution of household, family, social, community, and healthcare contacts to transmission. Dots represent empricial observations and violin plots represents model estimates based on 100 ensemble projections.

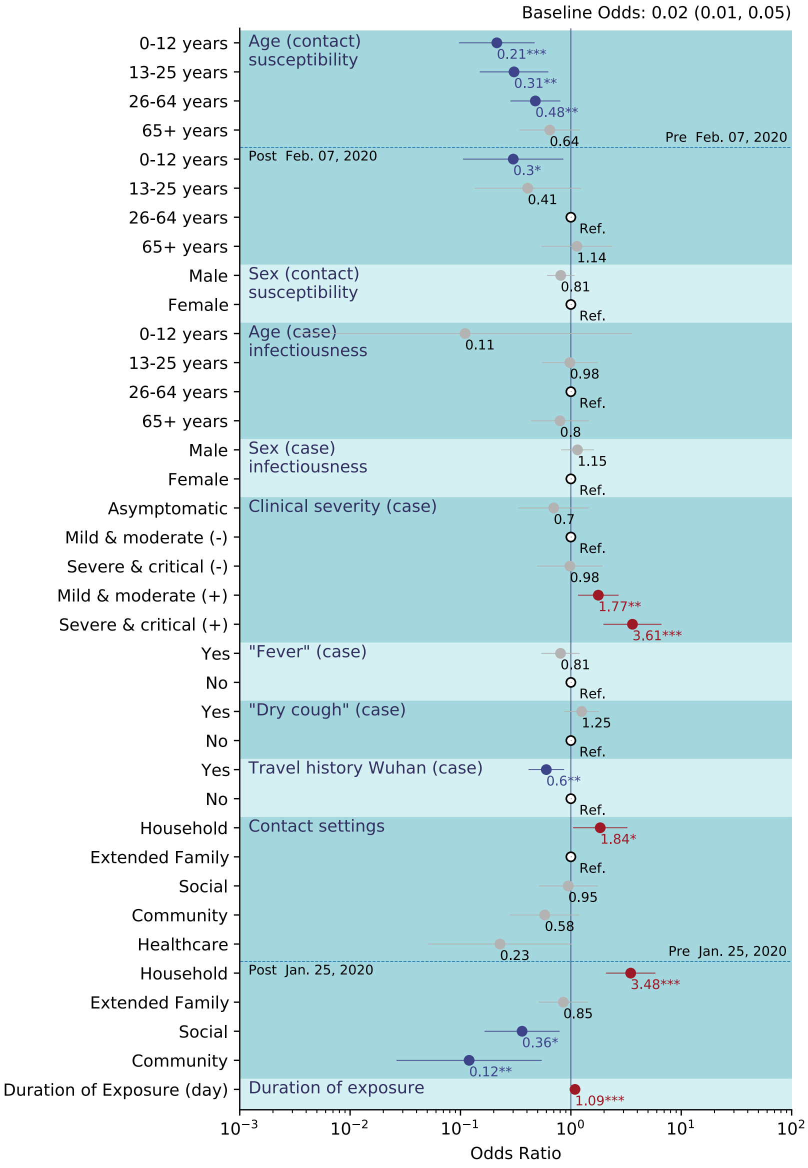


**Fig. S4.** A sensativity analysis of GLMM-logit regression that removes missing values and stratifies age of contacts by the date of change in testing protocols (February 7). The predictors of the logistic regression as those indicated on the left (fixed effects) and we also include random effects for individual SARS-CoV-2 infections. The "(-)" in "Mild & Moderate (-)" and "Sever & Critical (-)" indicate the SARS-CoV-2 infected individual's symptom onset occurred outside the exposure time window; The "(+)" in "Mild & Moderate (+)" and "Sever & Critical (+)" indicate the SARS-CoV-2 infected individual's symptom onset occurred within the exposure time window. Dots and lines indicate point estimates and 95% confidence interval of the odds ratio, numbers below the dots indicate the numerical value of the point estimates; "Ref." stands for reference category; * indicates p-value<0.05, ** indicates p-value<0.01, *** indicates p-value<0.001. Note that the regression results of odds ratio for healthcare contacts after January 25 is not visualized due to very low point estimate ($9.1 \times 10^{-8}$), with 0 of the 927 healthcare contacts after January 25 led to secondary transmissions.

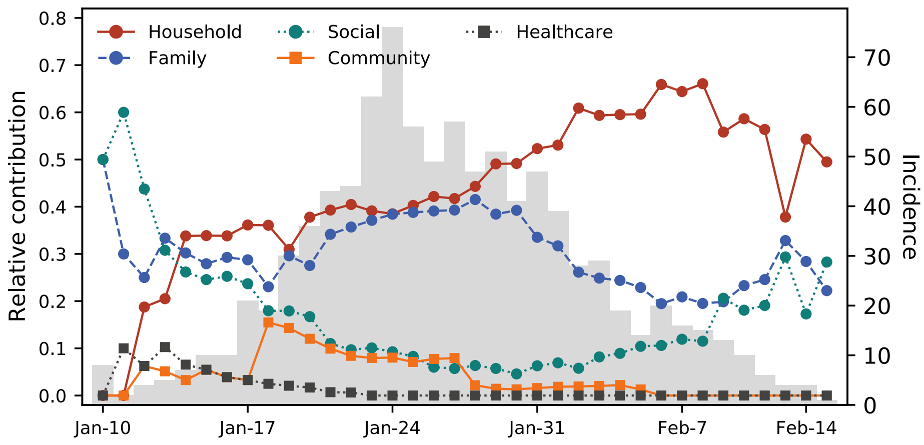


**Fig. S5.** Trends in the relative contribution of different types of contacts to SARS-CoV-2 transmission. Estimates are averaged over a 10-day moving window. The grey shade in the background indicates the time series of SARS-CoV-2 incidence in Hunan, China.

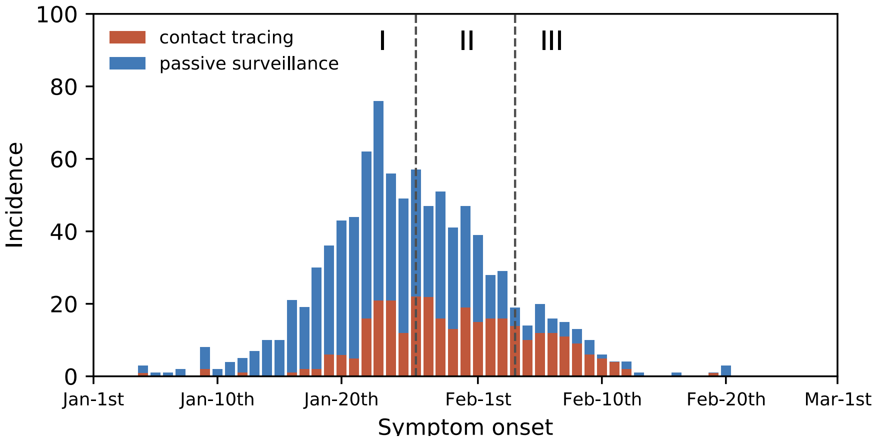


**Fig. S6**. Incidence of SARS-CoV-2 infections by onset date, for cases captured through contact tracing (red) or passive surveillance (blue). The dashes lines indicate the *Phase I, II, and III* of epidemic control.

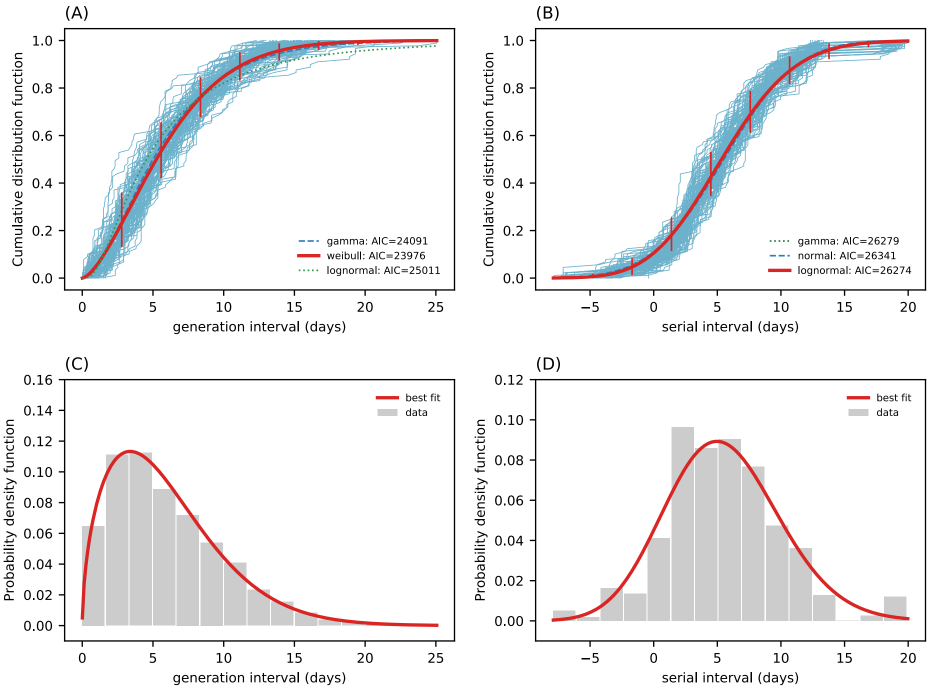


**Fig. S7.** (A) The cumulative distribution function of the generation interval distribution (time intervals between the infection of an infector and his/her infectee's). The thick blue solid lines represent each of the 100 realizations of the sampled transmission chains. The red solid line presents the best fit (Weibull distribution) to the ensemble average of the 100 realizations, based on the Akaike information criterion. Vertical lines represent the 95% confidence intervals of the Weibull distributions fitted to each of the 100 realizations individually. The vertical lines may not be visible due to narrow confidence intervals. Dashed lines represent lognormal and gamma distribution fitted to the data. (B) The cumulative distribution function of the serial interval distribution (intervals between the symptom onset time of an infector and his/her infectee's). The blue solid lines represent each of the 100 realizations of the sampled transmission chains. The solid red line presents the best fit distribution (lognormal distribution) to the ensemble average of the 100 realizations, based on the Akaike information criterion. Vertical lines represent the 95% confidence intervals of lognormal distributions fitted to each of the 100 realizations individually. The vertical lines may not be visible due to narrow confidence intervals. Dashed lines represent Weibull and gamma distribution fitted to the data. (C) and (D) visualize the probability density function of the best fit distribution (red line) for generation/serial intervals. The grey bars are histogram of generation/serial intervals with median and inter-quartile range of 5.3 (3.1, 8.6) days and 5.3 (2.7, 8.3) days respectively, based on the ensemble average of the 100 realizations.

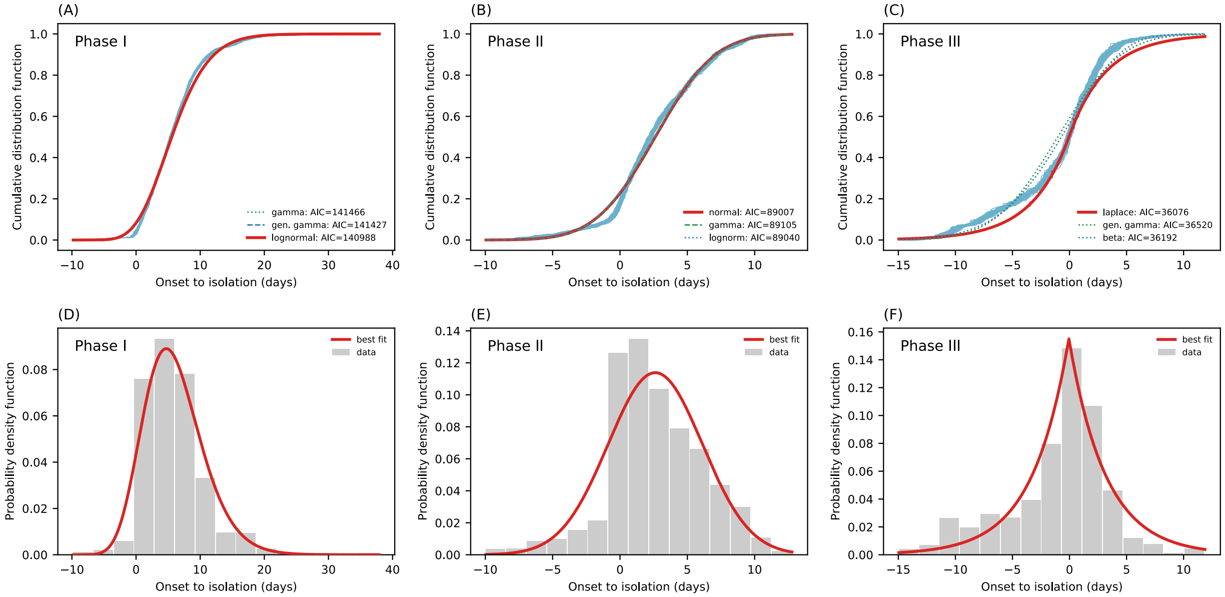


**Fig. S8.** Distribution of time from symptom onset to isolation in three different phases of epidemic control. Top row: The cumulative distribution functions of the onset to isolation distributions. The thick blue solid lines represent each of the 100 realizations of the sampled transmission chains. The thick solid red line presents the best fit to the ensemble average of the 100 realizations, based on the Akaike information criterion. Vertical lines represent the 95% confidence intervals of the best fit distribution to each of the 100 realizations individually. Noted that the dates of onset and isolation for each patient do not change across 100 realizations. However, the time point of onset/isolation of a patient was randomly sampled within the date of onset/isolation (1-day window). This will give rise to very moderate stochastic fluctuations of the onset to isolation distribution across 100 realizations. The vertical lines may not be visible due to narrow confidence intervals. Dashed lines represent alternative candidate distributions fitted to the data. Bottom row: Visualization of the probability density function of the best fit distribution (red line) for onset to isolation intervals. The grey bars are histogram of generation/serial intervals based on the ensemble average of the 100 realizations. (A) & (D), *Phase I* of epidemic control (before Jan. 27): time from onset to isolation has a median of 5.4 days with IQR (2.7, 8.2) days, based on the ensemble of 100 realizations of the sampled transmission chains. (B) & (E), *Phase II* of epidemic control (Jan. 27 – Feb. 4): time from onset to isolation distribution has a median of 2.2 days with IQR (0.4, 5.0) days, based on the ensemble of 100 realizations of the sampled transmission chains. (C) & (F), Phase III of epidemic control (after Feb. 4): time from onset to isolation distribution has a median of -0.1 days with IQR (-2.9, 1.8) days, based on the ensemble of 100 realizations of the sampled transmission chains.

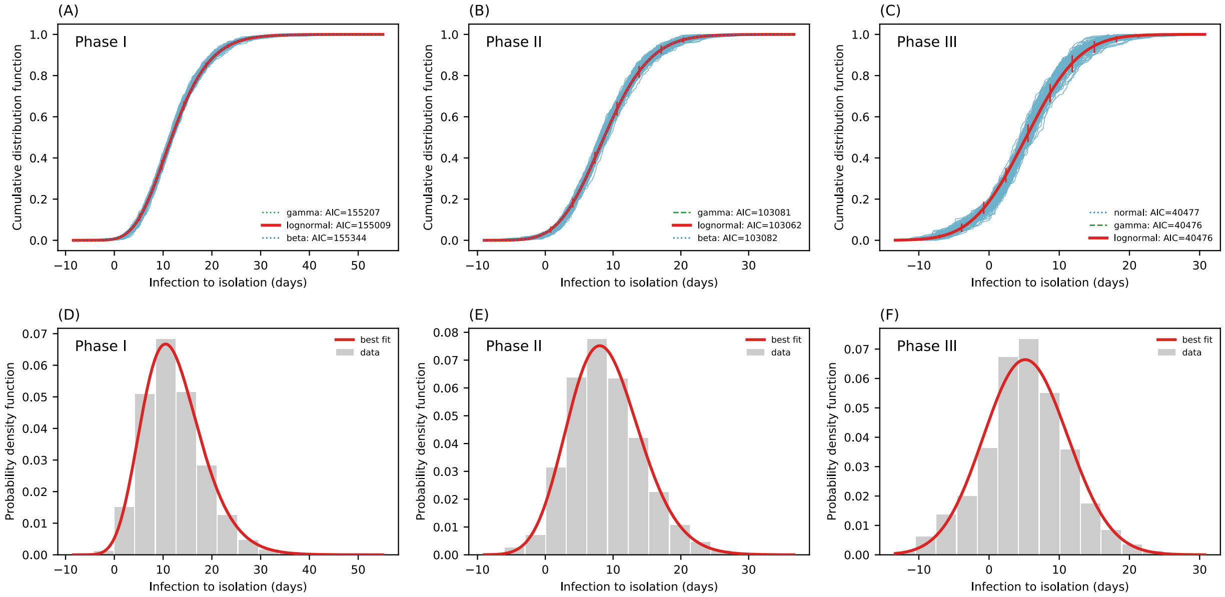


**Fig. S9.** Same as S8 but focusing on the time from infection to isolation. (A) & (D) Phase I of epidemic control (before Jan. 27): time from infection to isolation has a median of 11.6 days with IQR (8.0, 15.9) days (B) & (E) Phase II of epidemic control (Jan. 27 – Feb. 4): time from infection to isolation distribution has a median of 8.5 days with IQR (5.2, 12.3) days. (C) & (F) Phase III of epidemic control (after Feb. 4): time from infection to isolation distribution has a median of 5.4 days with IQR (1.9, 9.1) days.

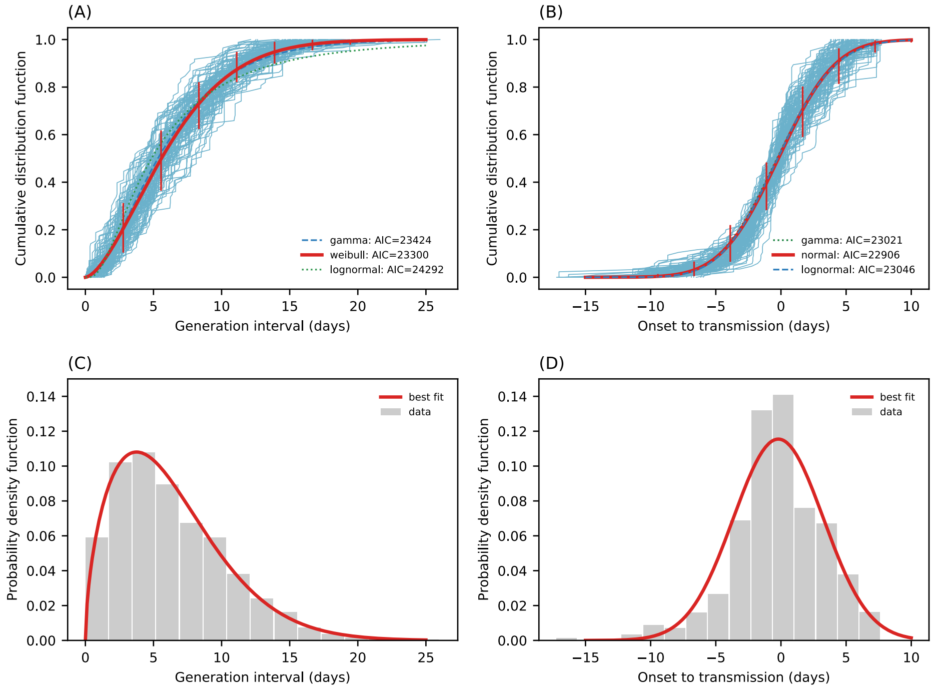


**Fig. S10.** (A) The cumulative distribution function of generation interval $\tau_{\mathrm{GI}}^{\mathrm{adj.}}$ distribution adjusted for censoring due to case isolation and quarantine. This represents the distribution that would have been observed in the absence of quarantine and case isolation. The blue solid lines represent each of the 100 realizations of the sampled transmission chains. The red solid line presents the best fit (Weibull distribution) to the ensemble average of the 100 realizations, based on the Akaike information criterion. Vertical lines represent the 95% confidence intervals of the Weibull distributions fitted to each of the 100 realizations individually. The vertical lines may not be visible due to narrow confidence intervals. Dashed lines represent lognormal and gamma distribution fitted to the data. (B) The cumulative distribution of time from symptom onset to transmission $\tau_{OT}^{adj.}$; negative values represent pre-symptomatic transmission. The blue solid lines represent each of the 100 realizations of the sampled transmission chains. The red solid line presents the best fit (normal distribution) to the ensemble average of the 100 realizations, based on the Akaike information criterion. Vertical lines represent the 95% confidence intervals of the normal distributions fitted to each of the 100 realizations individually. The vertical lines may not be visible due to narrow confidence intervals. Dashed lines represent lognormal and gamma distribution fitted to the data. (C) and (D) visualize the probability density function of the best fit distribution (red line) for $\tau_{\mathrm{GI}}^{\mathrm{adj.}}$ and $\tau_{OT}^{adj.}$ separately. The grey bars are histogram of $\tau_{\mathrm{GI}}^{\mathrm{adj.}}$ and $\tau_{OT}^{adj.}$, with median and interquartile range as 5.4 (3.1, 8.7) days and -0.2 (-2.0, 2.0) days respectively, based on the ensemble average of the 100 realizations.

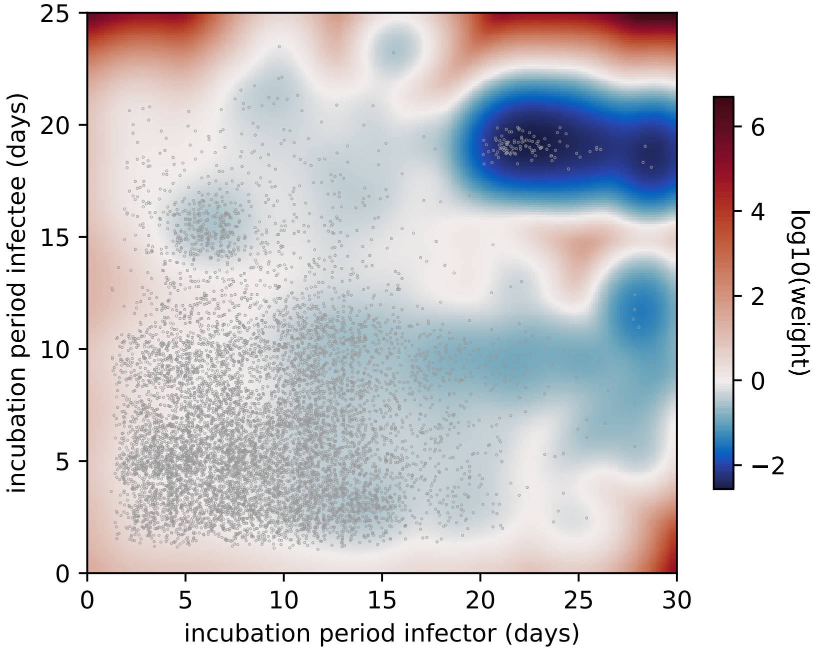


**Fig. S11.** Resampling weight of the incubation period pairs of infector/infectee. The heatmap visualizes the resampling weights of the incubation period pairs of infector/infectee. The blue region (resampling weight < 1) indicates down-sampling while the red region (resampling weight > 1) indicate over-sampling. The grey dots are incubation period pairs to be sampled. The cluster of outliers in the upper right corner is heavily down sampled.

**Table S1**. Definitions of clinical severity of SARS-CoV-2 infections

| **Clinical severity** | **Definition** |
|---|---|
| Asymptomatic | SARS-CoV-2 positive individuals who do not show any symptoms throughout the course of infection. |
| Mild | Patients with mild symptoms and no radiographic evidence of pneumonia |
| Moderate | Patients with fever, or respiratory symptoms, and radiographic evidence of pneumonia |
| Severe | Patients who have any of the following:<br>a. respiratory distress, breathing rate ≥30 beats/min; or<br>b. finger oxygen saturation ≤93% during resting state; or<br>c. $PaO_2/FiO_2$ ≤300mmHg (1mmHg = 0.133kPa).<br>Patients whose pulmonary imaging have obvious progress of lesions (>50%) within 24~48 hours are managed as severe case. |
| Critical | Patients who have any of the following:<br>a. respiratory failure that requires mechanical ventilation; or<br>b. shock; or<br>c. other organ failures that requires ICU admission. |

**Table S2:** Definition of contact types.

| **Contact Type** | **Definition** |
|---|---|
| Household | A household member living with a SARS-CoV-2 infected individual. |
| Extended family | A family member not residing in the same household but who has been in close contact with the primary SARS-CoV-2 infected individual. |
| Social | Friends, coworkers and classmates who study, work or are in close contact with the primary infected individual. |
| Community | Staff who interact with SARS-CoV-2-infected individuals in restaurants, entertainment venues, or other service settings; passengers seated in close proximity to a SARS-CoV-2 infected individual. |
| Healthcare | Healthcare workers who provide diagnosis, treat or nurse a SARS-CoV-2 patient or other patients and caregivers in the same ward as a SARS-CoV-2 infected individual. |

**Table S3:** Fixed effect variables of the mixed effects multiple logistic regression model

| Fixed effect | Definition | Category | Counts | % |
|---|---|---|---|---|
| Age (contact) | Age category of the contact. Age is categorized into three age categories: ***0-12 years***, ***13-25 years, 26-64 years***, ***65 years and older (+65 years)***; ***26-64 years*** is the reference category. | ***0-12 years*** | 1392/14662 | 9% |
| | | ***13-25 years*** | 1859/14662 | 13% |
| | | ***26-64 years*** | 9323/14662 | 63% |
| | | ***+65 years*** | 1415/14662 | 10% |
| | | ***NA*** | 673/14662 | 5% |
| Sex (contact) | Sex of the contact (***male/female***). ***Female*** is the reference category. | ***male*** | 7473/14662 | 51% |
| | | ***female*** | 6958/14662 | 47% |
| | | ***NA*** | 231/14662 | 2% |
| Age (case) | Age category of the case. Age is categorized into three age categories: ***0-12 years***, ***13-25 years, 26-64 years***, ***65 years and older (+65 years)***; ***26-64 years*** is the reference category. For main regression with data imputation (Fig. S3), we merge age brackets of ***0-12 years*** and ***13-25 years*** into ***0-25 years*** as only 3% of data in the ***0-12 years*** bracket. For regression of sensitivity analysis that removes missing data, we keep both ***0-12 years*** and ***13-25 years*** age brackets. | ***0-12 years*** | 27/870 | 3% |
| | | ***13-25 years*** | 74/870 | 8% |
| | | ***26-64 years*** | 666/870 | 77% |
| | | ***+65 years*** | 103/870 | 12% |
| | | ***NA*** | 0/870 | 0% |
| Sex (case) | Sex of the case (***male/female***). ***Female*** is the reference category. | ***male*** | 454/870 | 52% |
| | | ***female*** | 416/870 | 48% |
| | | ***NA*** | 0/870 | 0% |
| Clinical severity (case) | Clinical severity category of the case. Here we consider three categories: the first category represents ***asymptomatic*** cases; the second represents ***mild & moderate*** cases (reference category) and the third represents ***severe & critical*** cases. A definition of clinical severity is provided in Section 1.1, Table S1. | ***asymptomatic*** | 108/870 | 12% |
| | | ***mild*** | 217/870 | 25% |
| | | ***moderate*** | 427/870 | 49% |
| | | ***severe*** | 94/870 | 11% |
| | | ***critical*** | 24/870 | 3% |
| | | ***NA*** | 0/870 | 0% |
| "Fever" (case) | If the SARS-CoV-2 case had ***"fever (Yes/No)"*** during the course of illness. Cases without ***"fever"*** are the reference class. | ***Yes*** | 524/870 | 60% |
| | | ***No*** | 342/870 | 39% |
| | | ***NA*** | 4/870 | 1% |
| "Dry cough" (case) | If the SARS-CoV-2 case had ***"dry cough (Yes/No)"***. Cases without ***"dry cough"*** are the reference class. | ***Yes*** | 314/870 | 36% |
| | | ***No*** | 552/870 | 63% |
| | | ***NA*** | 4/870 | 1% |

| | | | | |
|---|---|---|---|---|
| Travel history Wuhan (case) | If the SARS-CoV-2 case had ***travel history to Wuhan (Yes/No)***: cases without ***travel history to Wuhan*** are the reference category. | ***Yes*** | 356/870 | 41% |
| | | ***No*** | 459/870 | 53% |
| | | ***NA*** | 55/870 | 6% |
| Contact types | The type of interactions between a case and a contact: the 5 contact types are ***household***, ***extended family***, ***social***, ***community***, and ***healthcare*** contacts. Definition of the contact types are detailed in Section 1.2 Table S2. For exposures of each contact type, we first define the exposure time as the midpoint between the start and end date of the exposure window. The median household exposure time is 01/25, 2020. We further divide each type of contacts into two categories: we denote household contact with exposure time before 01/25/2020 as ***household (pre 01/25)***; we denote household contact with exposure time after 01/25/2020 as ***household (post 01/25)***. The reference class is ***household (pre 01/25)***. Similarly, we divide ***extended family***, ***social***, ***community***, and ***healthcare*** contacts into: ***extended family (pre/post 01/25), social (pre/post 01/25), community (pre/post 01/25), healthcare (pre/post 01/25).*** | ***household (pre 01/25)*** | 964/17750 | 5% |
| | | ***household (post 01/25)*** | 924/17750 | 5% |
| | | ***extended family (pre 01/25)*** | 3141/17750 | 18% |
| | | ***extended family (post 01/25)*** | 2723/17750 | 15% |
| | | ***social (pre 01/25)*** | 2269/17750 | 13% |
| | | ***social (post 01/25)*** | 1626/17750 | 9% |
| | | ***community (pre 01/25)*** | 3328/17750 | 13% |
| | | ***community (post 01/25)*** | 822/17750 | 5% |
| | | ***healthcare (pre 01/25)*** | 740/17750 | 4% |
| | | ***healthcare (post 01/25)*** | 927/17750 | 5% |
| | | ***NA*** | 1313/17750 | 7% |
| Duration of exposure | Duration of exposure: defined as the time interval between the start and end date of exposure in days. The duration of exposure is a numeric variable. | ***NA*** | 2045/17750 | 11% |
| Onset within exposure | Onset within exposure: defined as if the symptom onset of the primary case occurred within the exposure time window of the contact. (***Yes/No***), ***No*** is the reference class. | ***Yes*** | 9837/17750 | 56% |
| | | ***No*** | 4118/17750 | 23% |
| | | ***NA*** | 3795/17750 | 21% |

**Table S4:** Sensitivity analysis of fitting alternative regression models with fewer contact type categories

| Model | Contact type categories | AIC |
|---|---|---|
| **M0** | The reference model, with contact types described as in Table S3, with 10 categories including: ***household (pre 01/25), household (post 01/25), extended family (pre 01/25), extended family (post 01/25), social (pre 01/25), social (post 01/25), community (pre 01/25), community (post 01/25), healthcare (pre 01/25), healthcare (post 01/25).*** | 2272 |
| **M1** | Compared to M0, we do not distinguish timing of exposure in this model and merge all contact of the same category, irrespective of time. We have 5 categories solely based on contact types, namely: ***household, extended family, social, community, healthcare.*** | 2274 |
| **M2** | Compared to M1, we further merge ***household*** and ***extended family*** contacts together with 4 contact categories, namely: (***household + extended family), social, community, healthcare.*** | 2323 |
| **M3** | Compared to M2, we further merge ***household, extended family, and social*** contacts together with 3 contact categories, namely: (***household + extended family + social), community, healthcare.*** | 2347 |
| **M4** | Compared to M3, we further merge ***household, extended family, social, and community*** contacts together with 2 contact categories, namely: (***household + extended family + social +community), healthcare.*** | 2365 |
| **M5** | We remove contact types as a predictor of the regression all together. | 2382 |
| **M6** | The contact types are the same as the baseline model (M0), however we introduce an interaction term between contact type and contact duration. | 2274 |

**Table S5:** Variables of the negative binomial regression on cumulative contact rates.

| Independent variable | Definition |
|---|---|
| Age (categorical) | Age category of the SARS-CoV-2 case. We consider three age categories: ***0-18 years***, ***19-64 years***, ***65 years and older***; ***19-64 years*** is the reference category. |
| Sex (Male/Female) | Sex of the contact (***male/female***). ***Female*** is the reference category. |
| Symptom fever (Y/N) | Whether the SARS-CoV-2 case had ***"fever"***. Cases without ***"fever"*** are the reference class. |
| Symptom dry cough (Y/N) | Whether the SARS-CoV-2 case had ***"dry cough"***. Cases without symptom ***"dry cough"***, i.e. Dry Cough (N), is the reference class. |
| Travel history to Wuhan (Y/N) | If the SARS-CoV-2 case reported a ***travel history to Wuhan***: cases without ***travel history to Wuhan*** are the reference category. |
| Physical distancing (Before/After Jan. 25) | Based on the within-city mobility index (Fig. S3A, insert) provided by Baidu Qianxi (*25*), we grouped the individual patients into categories depending on whether the patients symptom onsets occurred before and after January 25, 2020, corresponding to weak/strong physical distancing. Onsets occurred ***before Jan. 25*** (weak physical distancing) is the reference class. |
| Isolation to onset (days) | Time from case isolation to symptom onset. This is used as a proxy for individual-level intervention intensity. The larger the value, the earlier the case is being isolated. Positive values indicate isolation before symptom onset, negative values indicate isolation after symptom onset. |
